# Novel adenoma-immune phenotypes are associated with risk of metachronous polyps and colorectal cancer in a bowel screening cohort

**DOI:** 10.64898/2026.02.25.26346992

**Authors:** Stephen T McSorley, Tomoko Iwata, Aula Ammar, Sara S.F. Al-Badran, Lewis Irvine, Claire Kennedy Dietrich, Assya Legrini, Marjolein R de Koning, Natalie Fisher, Emma C Parsons, Philip Dunne, Calum Sampson, Gabriel Reines March, Noori Maka, Nigel B Jamieson, Mark Johnstone, Gerard Lynch, Joanne Edwards

## Abstract

**Background:** Current British Society of Gastroenterology (BSG) guidelines misclassify metachronous lesion risk after polypectomy in approximately 40% of patients. Building on evidence that immune exclusion drives progression of adenomas to colorectal cancer, this study examined immune profiles in screen-detected adenomas as a predictive biomarker for metachronous lesion risk.

**Methods:** Patients undergoing polypectomy within the Scottish Bowel Screening Programme, with surveillance colonoscopy between 6 months and 6 years were included (n=2642). Chromogenic immunohistochemistry (IHC; n=2466), 6-plex multiplex immunofluorescence (mIF; n=331), and spatially resolved 6000-plex single cell transcriptomics (n=7) were applied to adenoma microarrays. Cell density and location were measured using QuPath. Hierarchical then K-means clustering was used to define immune cell density-based clusters, which were compared to future lesion events using Kaplan-Meier curves and the log rank test.

**Results:** After adjustment for age, sex, site, size and dysplasia, adenoma CD3^+^ T cell density was significantly associated with future colorectal neoplasia (OR 1.44, 95% CI 1.20-1.72, p<0.001). Using mIF three immune cell density clusters were identified; 1) high T cell density, low macrophage density, 2) low T cell density, low macrophage density, and 3) high T cell, macrophage and αSMA density, with significant differences in future lesion risk (Cluster 1: 22%, Cluster 2: 41%, Cluster 3: 36%, *p*=0.032). Bulk RNAseq and spatial transcriptomic analysis revealed significant variation in T cell and macrophage co-location and gene expression profiles between clusters.

**Conclusion:** Adenoma immune contexture emerges as a possible determinant of future metachronous lesion risk, offering a novel biomarker to refine surveillance.

## Introduction

Approximately 30-40% of individuals undergoing screening colonoscopy will have pre-malignant colorectal polyps, of which the majority are conventional adenomas [1]. Endoscopic polypectomy may prevent future malignant transformation; however, patients remain at increased risk of going on to develop further polyps and/or colorectal cancer (CRC)[2].

Current post-polypectomy surveillance protocols rely on broad parameters including polyp size, number and dysplasia at the index procedure [3]. However, as data from the Integrated Technologies for Improved Polyp Surveillance (INCISE) collaborative project has demonstrated they commonly lead to suboptimal prediction of risk with around 40% of those deemed high risk having no subsequent polyp and 47% of those considered low risk found to have a subsequent lesion [4]. As such, there is a rationale for developing new biomarkers that more precisely stratify patients according to their risk of metachronous lesions, whilst simultaneously providing deeper insights into colorectal cancer aetiology and highlighting potential avenues for interception [5].

The immune contexture within adenomas and the colonic mucosa may influence future risk of polyps and cancer [6]. There is emerging evidence that immune exclusion, particularly of beneficial anti-cancer adaptive immune and cytotoxic effector responses, allows progression of adenomas to cancer [7]. In screen detected adenoma tissue, relationships between immune infiltrates, in particular T cell subsets, and histopathological features associated with advanced adenomas including size, dysplasia and villous architecture, have been reported by our group and others [8–9].

This study aimed to determine whether the immune architecture and/or specific immune features of adenomas were associated with development of metachronous polyps or CRC within 6 years of an index bowel screening colonoscopy by integrating comprehensive demographic, exposure, clinical, and pathological data in a large highly annotated cohort.

## Methods

### Patients

The INCISE cohort consists of 2642 bowel screening patients from NHS Greater Glasgow and Clyde (2009-2016) who had colonoscopic polypectomy and underwent at least one surveillance colonoscopy 6 months - 6 years after (Figure 1), with the primary outcome of metachronous polyp(s) or CRC during that period [4]. Recorded data included age, sex, medication, co-morbidities, some exposures (Clinical Portal, Orion Health, Boston USA; Trakcare, Intersystem, Boston USA), endoscopy reports (Unisoft GI Reporting Software, v2.5, Unisoft Medical Systems, UK), and histopathology results (Telepath Laboratory Information Management System). Serrated lineage lesions were excluded due to their prevalence, and the differing background immune infiltrates between these and conventional adenomas [9]. Patients with CRC, history of CRC, diagnosed polyposis or CRC predisposition syndromes, inflammatory bowel disease, or systemic vasculitis were excluded.

**Figure 1.**
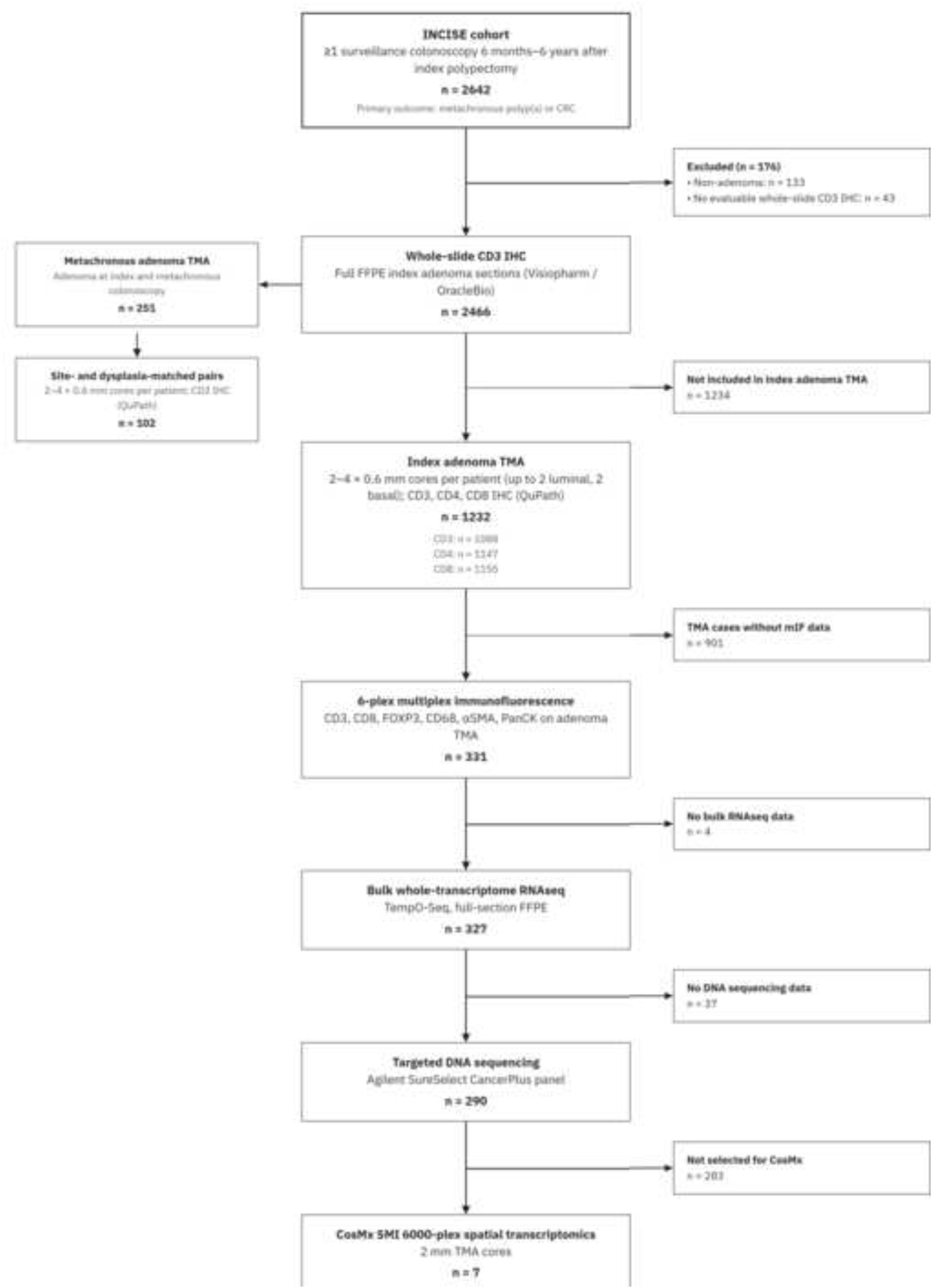
Patient and adenoma sample flow chart.

### Patient Tissue

Both 4-µm thick whole slide formalin-fixed and paraffin-embedded (FFPE) adenomas and 2.5-μm thick adenoma tissue microarrays (TMAs) were used. Whole slide images were re-reviewed by an NHS Consultant Histopathologist (NM) to confirm that they were conventional adenomas. TMAs were constructed using a TMA Grandmaster (3DHistech, Budapest, Hungary). Each patient within a TMA was represented by up to four 0.6 mm diameter cores. A pathologist identified and annotated areas encompassing superficial luminal facing- and basal crypt regions within an adenoma, with 2 cores from each.

### Immunohistochemistry

FFPE adenoma samples were stained with anti-CD3 (1:100, NCL-CD3-565, Leica Biosystems, UK), anti-CD4 (NCL-L-CD4-368, Leica Biosystems, UK), anti-CD8 (1:400, Clone C8/144B, Dako, Denmark), B-Catenin (1:1000, M3539, Dako, Denmark) and E-Cadherin (1:5000, 610182, BD Biosciences, USA) antibodies using the Ultravision Quanto Detection System HRP (Epredia, NH, USA). Whole slides were scanned at x40 and TMA slides at x20 resolution in the Glasgow Tissue Research Facility (GTRF) with a Hamamatsu S60 NanoZoomer (Hamamatsu, Japan) and images were stored within NZConnect (v 1.1.0) and accessed using NDP Viewer.

### Assessment of whole slide IHC Immune Cell Counts

Cell density (cells/mm^2^) was determined by point count of CD3^+^ cells in whole slide adenoma FFPE samples (n=2466) using Visiopharm apps (Visiopharm A/S, Hoersholm, Denmark) developed by OracleBio. The image analysis workflow was composed of four stages: (i) identification of viable tissue and major artefacts; (ii) tissue classification into adenoma and non-adenoma compartments; (iii) subdivision of adenoma/non-adenoma compartments into epithelium and lamina propria regions of interest (ROIs); and (iv) cell segmentation and classification (CD3^+^, CD3^-^) within each ROI (Figure 2C).

**Figure 2.**
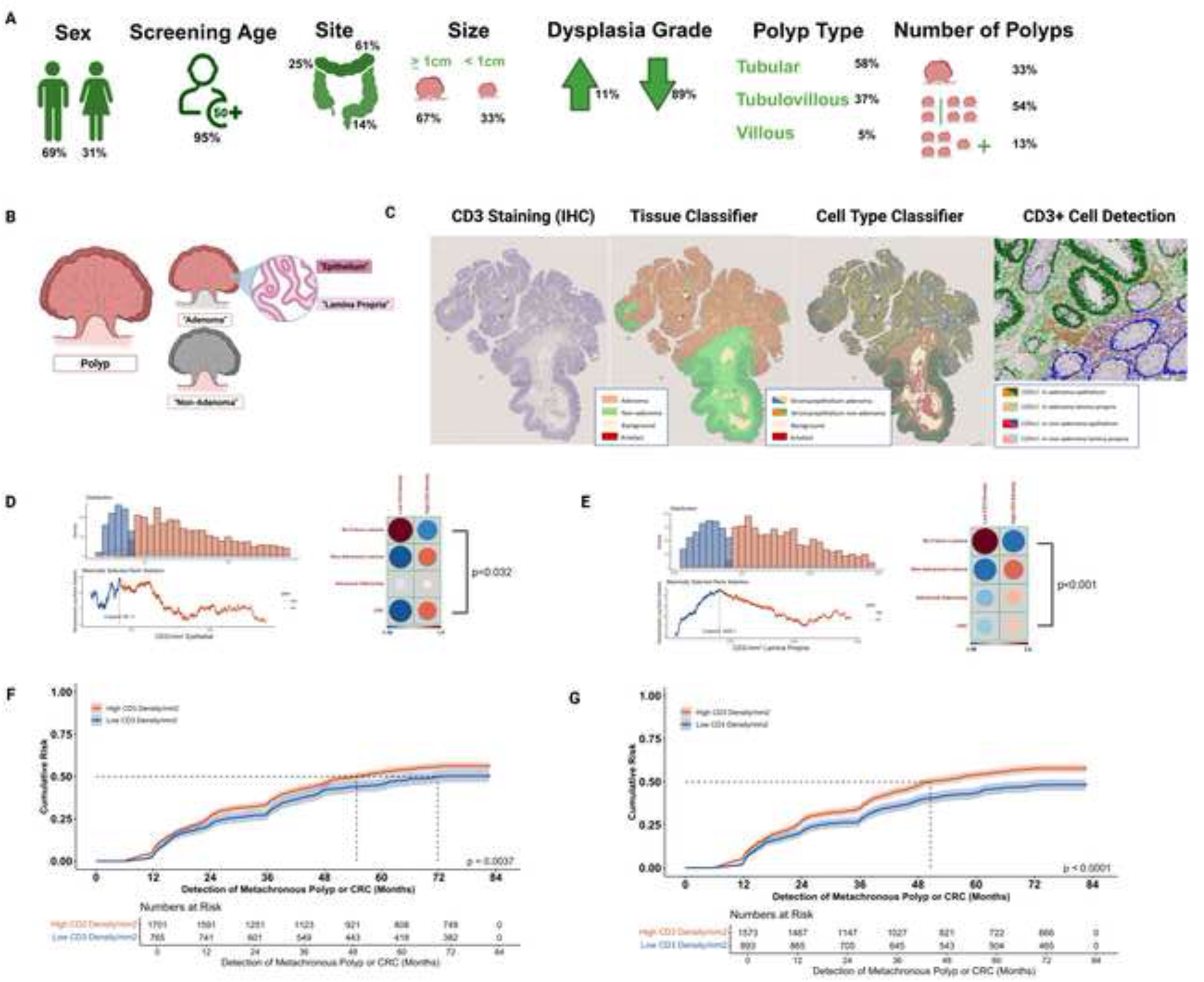
CD3^+^ Cell Density Investigated in Full Sections of Colorectal Adenomas. **A)** Description of clinical characteristics of patients and FFPE colonic adenomas removed at bowel screening polypectomy (n=2466) cohort. **B)** Schematic representing defined histological locations of the polyp. **C)** Full tissue sections were stained for CD3^+^ using chromogenic IHC, scanned to a digital pathology server and CD3^+^ density measured in the adenoma epithelium and lamina propria using a Visiopharm-based app developed by OracleBio following QC and tissue segmentation. After CD3^+^ density was dichotomised using the *survminer* R package, corrplots were generated to display the associations between CD3^+^ cell density and future lesion outcomes in the **D)** epithelium and **E)** lamina propria, with dot size and colour intensity (as directed by the z-score indicator) showing a significant correlation. **F)** Kaplan-Meier curves with log-rank statistics revealed high CD3^+^ density in the adenoma epithelium was significantly associated with higher risk of future polyp or colorectal cancer 6 months to 6 years after the index procedure (p=0.004). **G)** Kaplan-Meier curves with log-rank statistics revealed high CD3^+^ density in the adenoma lamina propria was significantly associated (p<0.0001) with higher risk of future polyp or colorectal cancer 6 months to 6 years after the index procedure.

CD3^+^ cell density within each tissue compartment in FFPE adenoma whole slides were dichotomised using *survminer* (Ver.0.4.9) and *Maxstat* (Ver. 0.7-25). SPSS (Ver. 28.0.1.1, IBM, NY, USA) was used to examine associations between CD3^+^ cell density and demographic, clinical and pathological characteristics using chi-squared tests for categorical data and Mann-Whitney tests for continuous data where *p*<0.05 was considered statistically significant.

Time to the event of future polyp or CRC during surveillance was analysed in relation to CD3^+^ T cell density groups using Kaplan-Meier analysis and the log rank test. CD3^+^ T cell density was entered into a multivariable binary logistic regression analysis model with sex, age, lesion site, size, dysplasia, histological subtype and total polyp number, with a statistical significance threshold of *p*<0.05 used to identify independent association with metachronous polyps or CRC.

Missing data were excluded on a variable by variable basis with no replacement or imputation.

### Assessment of TMA IHC

T cells were quantified on TMA using QuPath (Version 0.5.1)[10]. TMA images were de-arrayed into a grid and cores with inadequate quality were excluded. Positive cell detection determined the number of CD3^+^, CD4^+^ and CD8^+^ cells in each core. A Random Trees object classifier was trained using two annotations of polyp epithelium and lamina propria in a subset of cores, before applying to all other images. 10% of the images were manually scored and intraclass correlation coefficients (ICC) of >0.8 were achieved for all three markers. CD3^+^ cell density within each tissue compartment in FFPE adenoma whole slides and TMAs were compared by calculating Spearman’s correlation coefficient and Bland-Altman plot of paired mean vs difference. The density of CD3^+^, CD4^+^ and CD8^+^ T cell density in the adenoma epithelium and lamina propria were compared to clinical outcome using univariate time to event analysis as above. Weighted histoscores for nuclear B catenin and membranous E cadherin staining by epithelial and lamina propria compartment in epithelial cells were generated in QuPath. Around 10% of core were then assessed manually and weighted histoscores calculated [11], with ICCs >0.7 for both markers.

### Multiplex Immunofluorescence

Multiplex immunofluorescence (mIF) staining with antibodies against CD3, CD8, FOXP3, CD68, αSMA and Pan-cytokeratin (PanCK) was performed using Opal fluorophores and DAPI (Akoya Biosciences, Malborough, MA, USA). Slides were scanned using PhenoImager-HT (Akoya Biosciences). Following spectral unmixing by InForm, images were analysed using QuPath. Tissues and staining artefacts were visually inspected and excluded from ROI. Tissue segmentation into epithelium and lamina propria was performed by a pixel classifier generated by Random Trees algorithm using PanCK, αSMA and DAPI as detection channels. Object classifiers were generated and sequentially applied to detect CD3^+^, CD3^+^CD8^+^, CD3^+^FOXP3^+^ cells. CD68^+^ and αSMA^+^ cells were identified by cell detection using each channel. Cell density (cells/mm^2^) was calculated using number of detections divided by area of ROIs.

Hierarchical (using Ward’s method), then k means clustering identified and assigned membership of groups based on mIF immune cell density. Cluster centroids were compared using MANOVA. Cluster groups were compared to time to event outcome data as described above, then to clinicopathological, exposure, and prescribing data using the Chi-squared test with p<0.05 considered statistically significant.

### DNA Sequencing and Mutational Analysis

Sample sequencing and variant calling was performed by the Genomic Innovation Alliance (Glasgow, UK) using the Agilent SureSelect CancerPlus panel (Design ID: S3225252) and the quality and quantity of libraries were determined by TapeStation using a D1000 ScreenTape (5067-5582, Agilent, Santa Clara, CA). Variants were called using Genomic Innovation Alliance’s HOLMES pipeline.

Mutational analysis was performed on Rstudio (2023.12.0+369; MA, USA; R Core Team 2021) using *maftools* (Ver. 2.18.0) under the BiocManager Repository (Ver. 1.30.22). CoBar plots comparing the top 12 mutated genes between groups were generated using the “coBarplot” function. All statistical significance for mutational analysis was set at an adjusted p-value (padj) of <0.05.

### Bulk RNAseq

Whole human transcriptome bulk RNAseq was carried out on full-section FFPE adenoma samples by BioClavis Ltd (Biospyder Technologies, Carlsbad, CA, USA) using their proprietary TempO-Seq™ technology. All bulk transcriptomic analysis was performed using R (4.3.3) in RStudio. After quality control and pre-processing using *sapply, stats* (Ver 4.2.3) and *Combat_seq* (sva Ver 3.46.0) probe IDs were mapped to gene symbols with duplicated genes collapsed using *MaxMeans* (*WGCNA* Ver. 1.72-1). *DESeq2* (Ver. 1.42.1) was used for differential gene expression analysis, and volcano plots generated using *ggplot2* (Ver. 3.5.0). *MCPcounter* (v1.2.0) estimated the abundance of tissue infiltrating immune and stromal cell populations using gene expression.

### Single Cell Spatial transcriptomics

The CosMx Spatial Molecular Imager (SMI) 6000-plex single cell spatial transcriptomics workflow was applied to 7 adenoma cores of 2-mm diameter (5-µm thickness) from 7 patients. The probe panel comprised 6200 genes, including 20 negative control probes. The slide was processed on CosMx SMI, with 112 fields of view (FOV) over 16 imaging cycles. Acquired images were then decoded into RNA transcripts and mapped to a corresponding segmented cell. The raw files were exported from the AtoMx platform for analysis in R. Morphology-stained images were reviewed on *Napari* (v.0.4.17) for quality issues including tissue lifting, excessive blurring and cell segmentation. Additionally, cell segmentation was checked post cell type annotation using *FastReseg* (v.1.0.2), only ∼2% of cells were flagged, confirming segmentation quality. The final round of QC filtered data based on count (>20), features (>20), size (>300um^2^) and negative proportion (<0.1) metrics. Final metrics were 112 FOVs, 147,449 cells/156,649 original cells and mean count per cell= 263.47.

Log normalization, variable features (selection method = vst, nfeatures = 2000) and scaling of data was all performed using *Seurat* (v.5.1.0). Batch correction using *harmony* (v.1.2.4) was run with Patient ID as the grouping variable.

Cell types were identified using semi-supervised methods in *InsituType* (v.2.0). The CosMx-single-cell Human Immune-Oncology (IO) 1K profile was used to identify the major immune and stromal cell types, with 5 unknown clusters generated including epithelial. Cell type density per core and cluster was compared across the mIF cluster groups.

Distance metrics were explored between CD68^+^ Macrophages, CD4^+^ T cells, CD8^+^ T cells and Tregs using *spatialTIME* (v.1.3.4-5). Differential expression analysis was performed using *smiDE* (v.0.0.2.04) comparing mIF cluster per cell type.

### Ethical approvals

Ethical approval was obtained for data (GSH/20/CO/002) and tissue analysis (22/WS/0020) from Glasgow SafeHaven, NHS Greater Glasgow and Clyde and the West of Scotland Research Ethics Committee.

## Results

### CD3^+^ cell density in the index adenoma is associated with metachronous polyp or CRC during post-polypectomy surveillance

Following tissue segmentation and detection of CD3^+^ cells in 2466 index adenoma FFPE full sections (Figure 2A-C), CD3^+^ cell density was calculated in the adenoma epithelium and lamina propria. CD3^+^ cell density in both compartments was strongly and significantly positively correlated (r_s_=0.728, *p*<0.001), with the median CD3^+^ density lower in the adenoma epithelium (121/mm^2^, IQR 75-186) than the lamina propria (2171/mm^2^, IQR 1554-2948). CD3^+^ cell densities were dichotomised, giving low (≤85/mm^2^, n=765) and high (>85/mm^2^, n=1701) density groups in the adenoma epithelium and low (≤1830/mm^2^, n=893) and high (>1830/mm^2^, n=1573) density groups in the lamina propria (Figure 2D). CD3^+^ density in the adenoma epithelium and lamina propria was associated with adenoma size (both *p*<0.001), and site (both *p*<0.001), while only CD3^+^ density in the adenoma lamina propria was associated with histological subtype (Table 1).

**Table 1:**
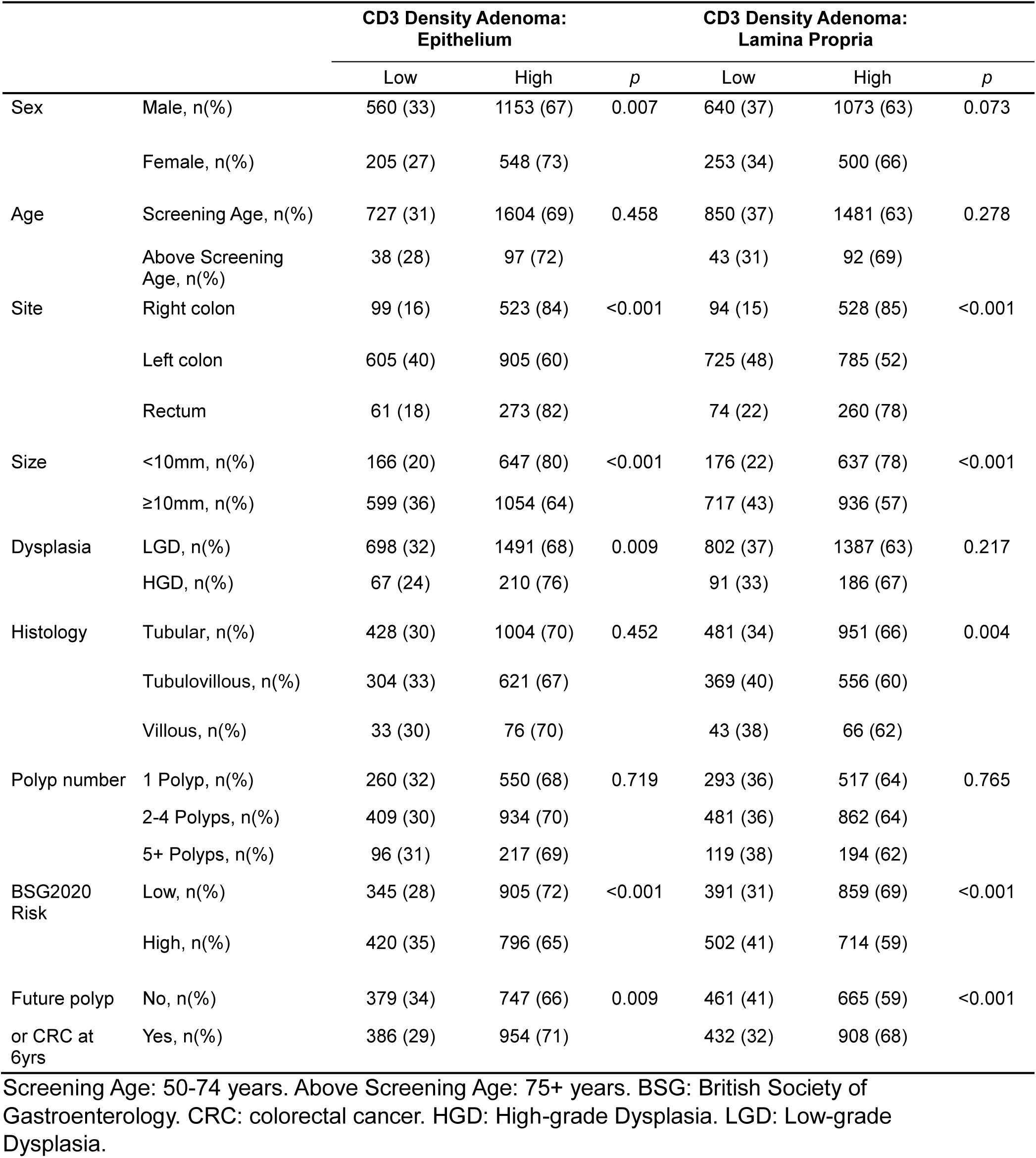
Patient and Index Adenoma Characteristics Grouped by CD3^+^ Cell Density in the Index Adenoma Epithelial and Lamina Propria Compartments.

|  |  | CD3 Density Adenoma:<br>Epithelium |  |  | CD3 Density Adenoma:<br>Lamina Propria |  |  |
| --- | --- | --- | --- | --- | --- | --- | --- |
|  |  | Low | High | *p* | Low | High | *p* |
| Sex | Male, n(%) | 560 (33) | 1153 (67) | 0.007 | 640 (37) | 1073 (63) | 0.073 |
|  | Female, n(%) | 205 (27) | 548 (73) |  | 253 (34) | 500 (66) |  |
| Age | Screening Age, n(%) | 727 (31) | 1604 (69) | 0.458 | 850 (37) | 1481 (63) | 0.278 |
|  | Above Screening Age, n(%) | 38 (28) | 97 (72) |  | 43 (31) | 92 (69) |  |
| Site | Right colon | 99 (16) | 523 (84) | <0.001 | 94 (15) | 528 (85) | <0.001 |
|  | Left colon | 605 (40) | 905 (60) |  | 725 (48) | 785 (52) |  |
|  | Rectum | 61 (18) | 273 (82) |  | 74 (22) | 260 (78) |  |
| Size | <10mm, n(%) | 166 (20) | 647 (80) | <0.001 | 176 (22) | 637 (78) | <0.001 |
|  | ≥10mm, n(%) | 599 (36) | 1054 (64) |  | 717 (43) | 936 (57) |  |
| Dysplasia | LGD, n(%) | 698 (32) | 1491 (68) | 0.009 | 802 (37) | 1387 (63) | 0.217 |
|  | HGD, n(%) | 67 (24) | 210 (76) |  | 91 (33) | 186 (67) |  |
| Histology | Tubular, n(%) | 428 (30) | 1004 (70) | 0.452 | 481 (34) | 951 (66) | 0.004 |
|  | Tubulovillous, n(%) | 304 (33) | 621 (67) |  | 369 (40) | 556 (60) |  |
|  | Villous, n(%) | 33 (30) | 76 (70) |  | 43 (38) | 66 (62) |  |
| Polyp number | 1 Polyp, n(%) | 260 (32) | 550 (68) | 0.719 | 293 (36) | 517 (64) | 0.765 |
|  | 2-4 Polyps, n(%) | 409 (30) | 934 (70) |  | 481 (36) | 862 (64) |  |
|  | 5+ Polyps, n(%) | 96 (31) | 217 (69) |  | 119 (38) | 194 (62) |  |
| BSG2020 Risk | Low, n(%) | 345 (28) | 905 (72) | <0.001 | 391 (31) | 859 (69) | <0.001 |
|  | High, n(%) | 420 (35) | 796 (65) |  | 502 (41) | 714 (59) |  |
| Future polyp or CRC at 6yrs | No, n(%) | 379 (34) | 747 (66) | 0.009 | 461 (41) | 665 (59) | <0.001 |
|  | Yes, n(%) | 386 (29) | 954 (71) |  | 432 (32) | 908 (68) |  |

Screening Age: 50-74 years. Above Screening Age: 75+ years. BSG: British Society of Gastroenterology. CRC: colorectal cancer. HGD: High-grade Dysplasia. LGD: Low-grade Dysplasia.

Significantly more patients were found to have metachronous polyps or CRC during surveillance in the high CD3^+^ density adenoma epithelium (p=0.004, Figure 2E) and lamina propria groups (*p*<0.001, Figure 2F) when compared to the low CD3^+^ density groups. On multivariate analysis (Table 2), accounting for sex, age, lesion site, size, dysplasia, histological subtype and total polyp number, CD3^+^ density remained significantly associated with risk of future polyp or CRC independent of these high risk patient and adenoma characteristics in both the adenoma epithelium (OR 1.21, 95% CI 1.01-1.45, *p*=0.042) and lamina propria (OR 1.44, 95% CI 1.20-1.72, *p*<0.001).

**Table 2:**
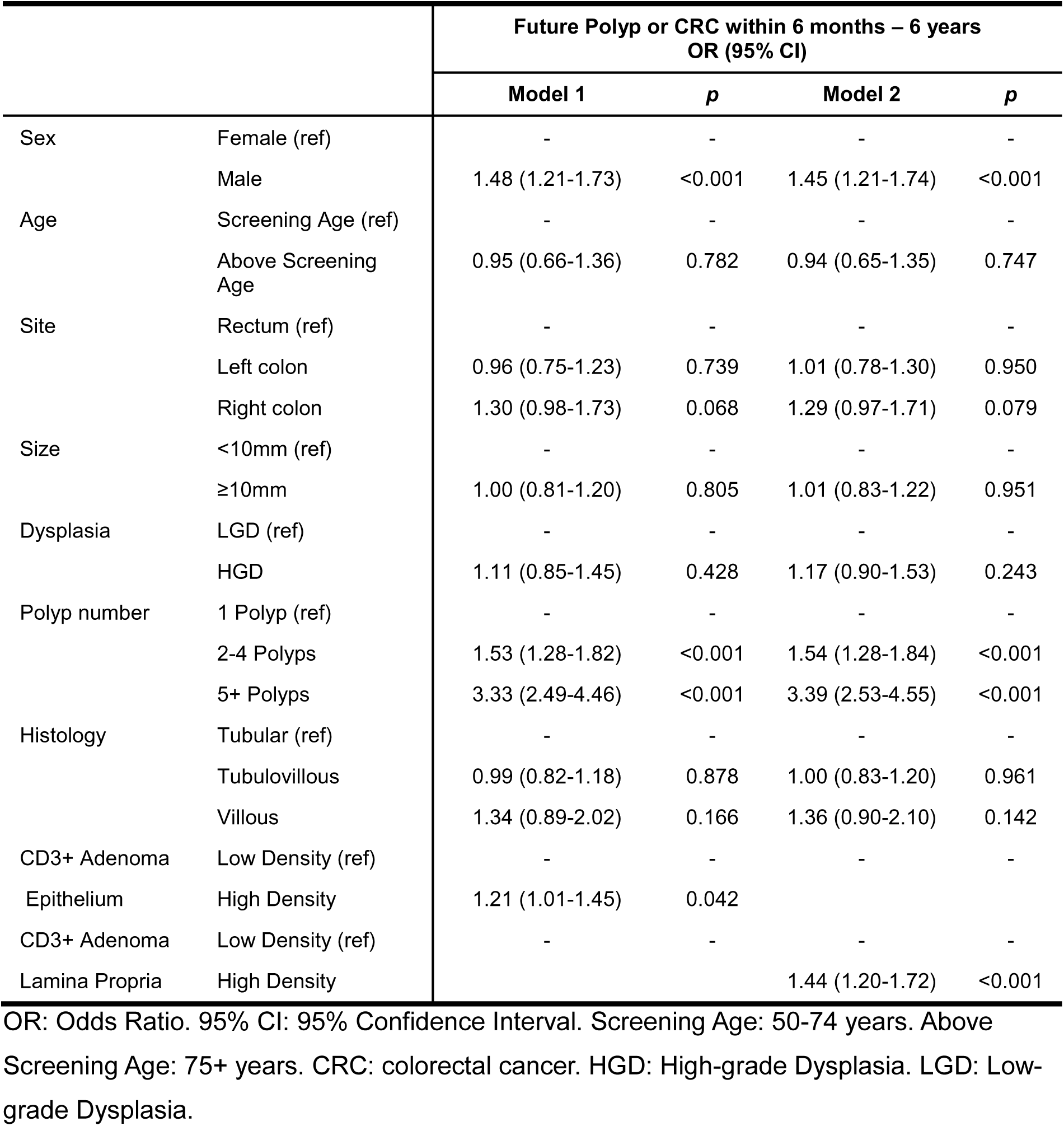
Multivariate Binary Logistic Regression Models of Factors Associated with Future Polyp or CRC During 6-Year Post-Polypectomy Surveillance Including Index Adenoma CD3^+^ Density in Adenoma Epithelium and Lamina Propria, in Bowel Screening Programme Polypectomy Patients.

### T cell density in adenoma TMAs recapitulates full section findings

TMAs were constructed for a sub-cohort of index adenomas (Figure 3A), capturing spatial heterogeneity by sampling up to four cores from across luminal and basal regions (Figure 3B). To confirm that TMAs provide a reliable platform for immune cell assessment, we repeated CD3⁺ IHC (Figure 3B). The comparable staining supported the validity of TMAs for immune profiling (r_s_=0.477, p<0.001)(Figure 3C–D). CD3⁺ positive cell counts were dichotomised (Supplementary Figure 1), with high CD3^+^ density in the adenoma epithelium (p=0.0018) and lamina propria (p=0.011) associated with shorter time to detection of metachronous lesion (Figure 3E). The TMAs were then sectioned and IHC performed for CD4^+^ and CD8^+^ cells, and the density of each in the adenoma epithelium and lamina propria dichotomised with *survminer* (Supplementary Figure 1). High CD4^+^ density in the adenoma epithelium (p=0.005) and lamina propria (p<0.001) was associated with shorter time to detection of metachronous lesion (Figure 3F), whereas CD8^+^ adenoma epithelium (p=0.078) and lamina propria (p=0.2) density was not (Figure 3G).

**Figure 3:**
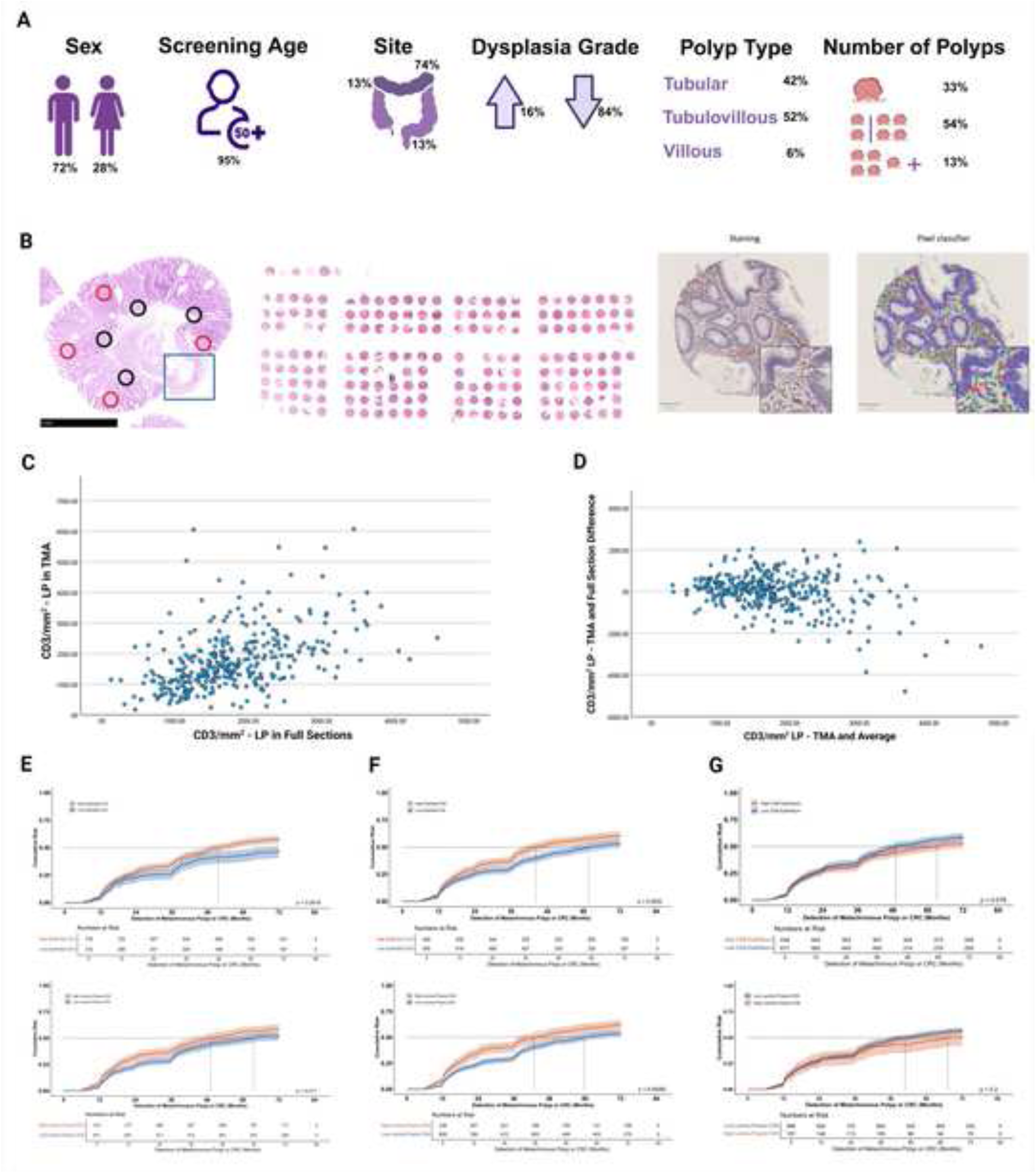
T Cell Densities in Adenoma TMAs Recapitulate Full Section Findings. **A)** Description of clinical characteristics of TMA cohort of FFPE colonic adenomas removed at bowel screening polypectomy (n=1232) cohort. **B)** Example of polyp areas cored to capture full tissue heterogeneity, with an example of a TMA slide. Red circles depict luminal areas and black circles depict basal areas (Left). Example of stained and digitally scored TMA core (Right). **C)** Scatter plot of CD3^+^ cell density in TMA lamina propria was significantly correlated with that in whole slide sections (r=0.477, *p*<0.001). **D)** Bland-Altman plot of mean against difference of full section and TMA lamina propria CD3^+^ density/mm^2^. **E)** CD3^+^ T cell counts were dichotomised, and higher CD3^+^ T cell in the epithelium (Top; *p*=0.0018) and in the lamina propria (Bottom; *p*=0.011) were significantly associated with higher risk of metachronous lesion. **F)** CD4^+^ T cell counts were dichotomised, and higher CD4+ T cell in the epithelium (Top; *p*=0.005) and in the lamina propria (Bottom; *p*<0.001) were significantly associated with higher risk of metachronous lesion. **G)** CD8^+^ T cell counts were dichotomised, and CD8^+^ T cell density was not associated with risk of metachronous lesion in either the epithelium (Top; *p*=0.078) or the lamina propria (Bottom; *p*=0.2).

In a further subcohort of patients (n=102) in whom a TMA had been created from a metachronous adenoma, matched to their paired index lesion by anatomic site and dysplasia grade (Supplementary Figure 2), CD3^+^ density in the lamina propria was positively correlated between the index and metachronous adenomas (Spearman’s rho=0.303, p=0.002).

### Clustering of immune cell density identified by mIF in adenoma TMAs reveals phenotypes associated with likelihood of future polyp or CRC

A 6-plex mIF panel was used to identify adaptive and innate immune populations in the epithelium and lamina propria in the adenoma TMA (n=331; Figure 4A and 4B). Hierarchical then k means clustering based on cell densities of CD3^+^, CD8^+^, FOXP3^+^, αSMA^+^ and CD68^+^ cells in the lamina propria produced 3 groups (Figure 4C). Cluster 1 had a mix of high T cell (CD3^+^, CD3^+^CD8^+^ and CD3^+^FOXP3^+^) density with low CD68^+^ and low αSMA^+^ density. In contrast, Cluster 2 had low T cell density, low CD68^+^ and low αSMA^+^ density, with a globally immune excluded phenotype. Finally, Cluster 3 showed high T cell, high CD68^+^ and high αSMA^+^ density.

**Figure 4:**
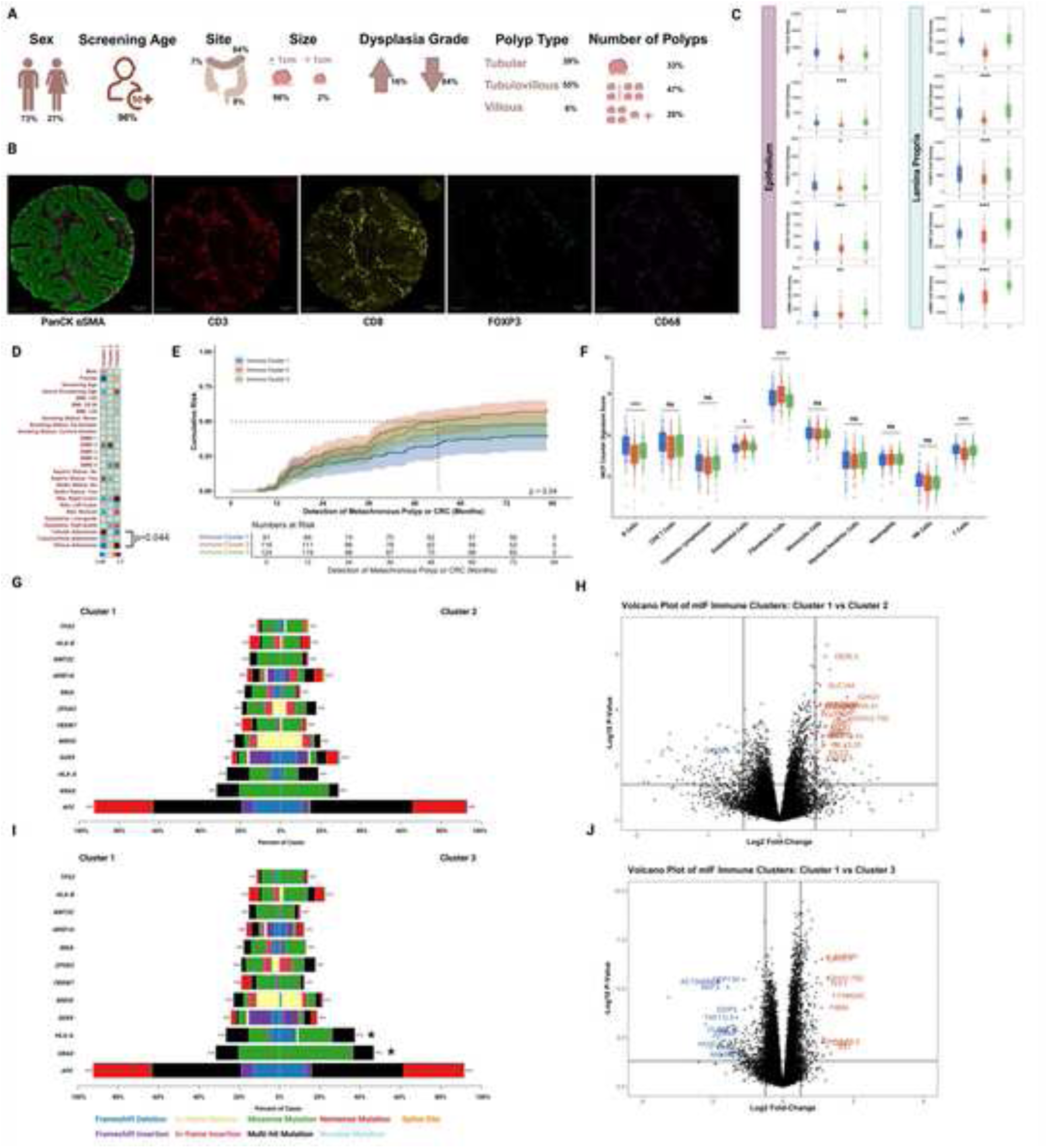
Immune Cell Densities Identified by mIF Reveal Predictive Phenotypes, Mutational Signatures and Differential Gene Expression. **A)** Description of the TMA sub cohort on which mIF was performed (n=331). **B)** 6-plex mIF immune panel of Pan-cytokeratin (PanCK), αSMA, CD3, CD8, FOXP3, and CD68 was visualised with PhenoImager-HT, with tissue segmentation and cell density measurement in QuPath. **C)** Hierarchical then k means clustering suggests three main immune clusters; Cluster 1 (blue) having a high T cell infiltrate with low macrophage and fibroblast density (“adaptive” infiltrate) in the lamina propria, Cluster 2 (red) having low density of all markers (“immune cold”) in the lamina propria, and Cluster 3 (green) having high density for all markers in the lamina propria (“mixed” infiltrate). **D)** Corrplot of the associations between clusters and patient, adenoma, or exposure characteristics. Dot size and colour intensity (as directed by the z-score indicator) suggests a high correlation between variables. **E)** Kaplan-Meier curve showing significant differences in future lesion risk between the three clusters (*p*=0.040) with the “adaptive” Cluster 1 having the lowest risk and the “immune cold” Cluster 2 having the highest risk. **F)** Bulk RNAseq of the FFPE adenomas analysed using Microenvironment cell populations (MCP)-Counter (v1.2.0), which estimated the abundance of tissue-infiltrating immune and stromal cell populations using gene expression. Boxplots show distribution of cell type signature scores across the 3 mIF derived adenoma-immune clusters, with significant differences in T cell, B cell and fibroblast signatures between clusters. **G)** Cobar plot of the top 12 mutated genes in index adenomas comparing Cluster 1 to Cluster 2. Genes are given in the y axis with the x axis denoting the proportion of samples within a cluster with at least 1 mutation called in that gene. There were no significant differences between Cluster 1 and Cluster 2. **H)** Volcano plot of relative gene expression with log2 fold change displayed on the x axis and -log10 p value on the y axis, comparing Cluster 2 (Right-hand side) to Cluster 1 (Left-hand side, baseline). **I)** Cobar plot of the top 12 mutated genes in index adenomas Cluster 1 to Cluster 3. Genes are given in the y axis with the x axis denoting the proportion of samples within a cluster with at least 1 mutation called in that gene. There were significantly higher proportion of adenomas with HLA-A and KRAS mutation in Cluster 3. **J)** Volcano plot of relative gene expression with log2 fold change displayed on the x axis and -log10 *p* value on the y axis, comparing Cluster 3 (Right-hand side) to Cluster 1 (Left-hand side, baseline). *Colour denotes mutation type for G and I. * denotes p<0.05, ** denotes p<0.01, and *** denotes p<0.001*.

No significant associations between the clusters and established risk factors, including age, sex, lesion histology, size, site, dysplasia, aspirin or statin prescription, smoking, or BMI were found. However, polyp histology was significantly associated with the immune clusters (*p*=0.044; Figure 4D, Supplementary Table 1).

The proportion of patients that were found to have a metachronous polyp or CRC during follow up were significantly different in each cluster (Cluster 1: 22%, Cluster 2: 41%, Cluster 3: 36%, *p*=0.032), which remained the case at time to event analysis (Figure 4E, *p*=0.040).

### Mutational signatures and bulk transcriptomic data validate mIF adenoma-immune clusters

Mutational data from Agilent’s Cancer Plus panel showed no significant differences in the frequency of the top 12 mutated genes between Cluster 1 and Cluster 2 (Figure 4G).

However, there were significant differences between Cluster 1 and Cluster 3 (Figure 4I) in the proportion of samples with at least one mutation in HLA-A (27% vs 37%, *padj*<0.05) and KRAS (32% vs 47%, *padj*<0.05), in keeping with a more immune suppressed or excluded environment [12]. IHC showed significantly higher (p=0.006) staining intensity for nuclear B catenin (Supplementary Figure 3) in Cluster 2 (median H-score=131), than either Cluster 1 (median H-score=117) or Cluster 3 (median H-score=103). There was no significant difference in H-score for membrane E-cadherin between clusters (p=0.507).

Bulk whole human transcriptome was examined in the context of mIF derived adenoma-immune clusters by TempO-Seq assay. A relative overexpression of genes associated with immunoglobulins, such as *IGHG1*, *IGLC2,* and *IGLV3.25,* was observed in Cluster 2 when compared to Cluster 1. Similarly, genes with family members associated with immune-tumour interaction roles and poorer prognosis in CRC and gastric cancers, like *POU2AF1* and *DERL3* [13–14], were upregulated in Cluster 2 when compared to Cluster 1 (Figure 4H). Cluster 3 also showed an overexpression of immunoglobulin genes (*IGKV3.7*, *IGHV2.70D,* and *IGHV1.69.2*) when compared to Cluster 1 (Figure 4J).

*MCPcounter* estimated the relative abundance of tissue-infiltrating immune and stromal cell populations by gene expression (Figure 4F). T-cell gene expression signature scored significantly higher in Cluster 1 than Cluster 2 or 3 (*p*<0.001), however there were no significant differences in CD8 or cytotoxic T-cell gene signature scores. The fibroblast gene signature score was significantly higher in Cluster 2 compared to Cluster 1 or Cluster 3 (*p*<0.001). This was discrepant to the αSMA mIF cluster centroid, however, it may represent the inability of bulk RNAseq to reflect variation in the tissue context of the epithelium and lamina propria across samples [15], or the presence of non αSMA-expressing myofibroblasts or pro-inflammatory fibroblasts in this group.

### Spatially resolved single cell gene expression signatures validate mIF adenoma-immune clusters and reveal adaptive immune exclusion based on macrophage phenotype and T cell interaction

Seven adenoma TMA samples from Cluster 1 (n=4), Cluster 2 (n=1) and Cluster 3 (n=2) underwent 6000-plex single cell spatial transcriptomics assessment using CosMx SMI (Figure 5A). UMAPs of single cell gene expression demonstrated clustering with respect to the three immune cell density-based groups previously defined by mIF (Figure 5B).

**Figure 5:**
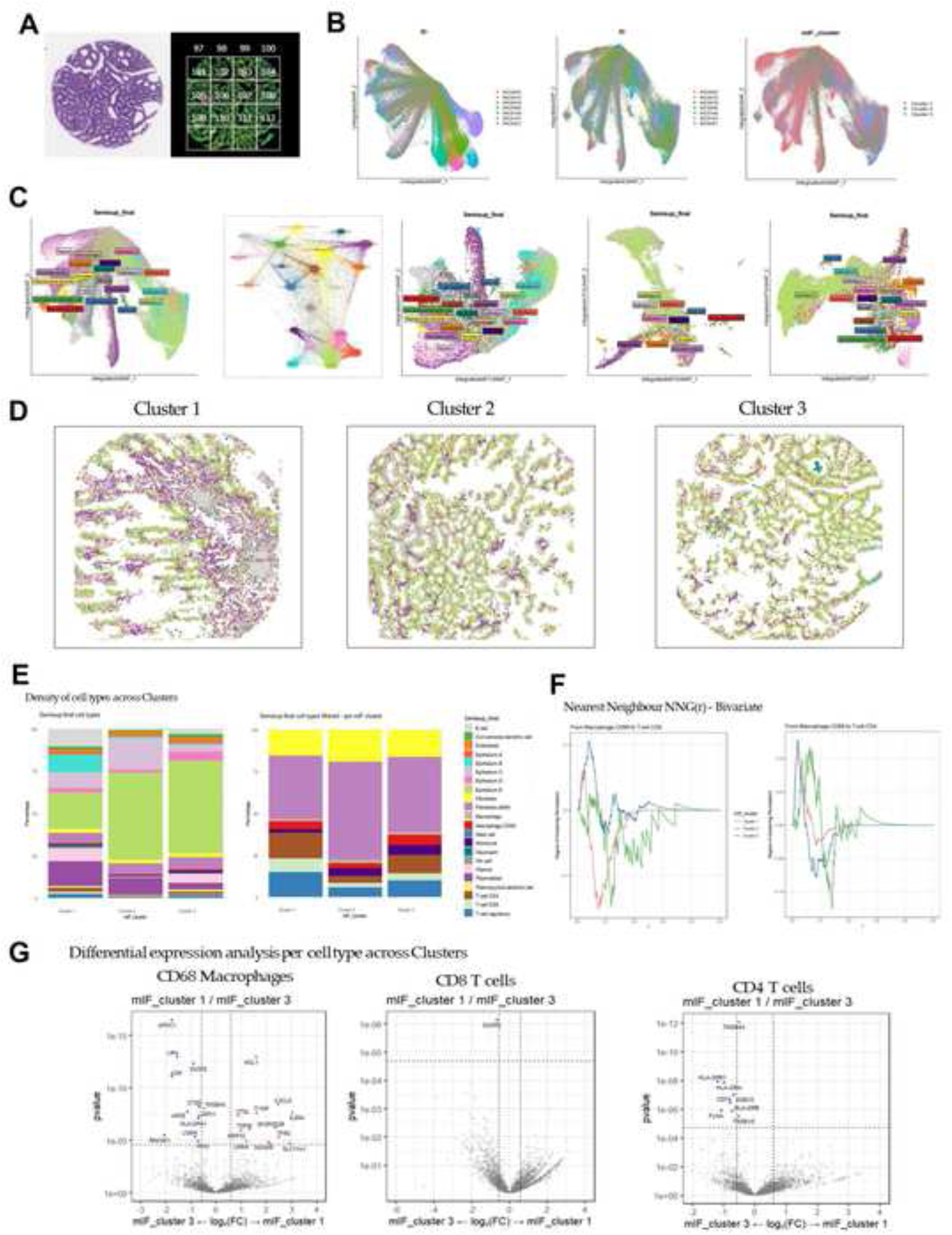
Spatial transcriptomic analysis of adenoma TMA cores reveals varying T cell and macrophage dynamics between mIF cluster groups. **A)** Adenoma TMA samples (n=7) underwent 6000-plex spatially resolved single cell RNAseq using CosMx SMI. **B)** Unintegrated and integrated UMAPs grouped by patient and the three predefined immune cell density clusters. **C)** UMAPs of cell type define relevant immune cell populations and five epithelial clusters across the three predefined immune cell density clusters. **D)** Cell typing overlay for a representative adenoma core from each of the 3 previously defined immune density clusters reflect the cell typing UMAP data in relation to variation in lamina propria immune cell density. **E)** Stack plots of proportions of cell type within each previously defined cluster show that Cluster 1 has the lowest proportion of epithelial cells and greater variety of different epithelial cell subtypes when compared to Cluster 2 and 3. Cluster 1 has the greatest proportion of T and B cells whilst Cluster 2 has the lowest, although the relative proportions of T cell subtypes are similar across the clusters. **F)** Nearest neighbour analysis suggests a pronounced short-range spatial association between CD68^+^ macrophages and CD8^+^ T cells in Cluster 3, while in comparison, Cluster 1 has predominant spatial separation between these cell types. **G)** There was little differential gene expression when CD8^+^ T cells were compared between Cluster 1 and Cluster 3, however CD4^+^ T cells in Cluster 3 overexpressed genes associated with antigen presentation and MHC II activity (HLA-DPB1, HLA-DRA, CD74) in comparison to those in Cluster 1 adenomas, while CD68^+^ macrophages in both clusters over-express genes associated with an immune-suppressive or pro-tumour microenvironment when expressed by TAMs in established solid cancers.

Following gene expression-based cell typing (Figure 5C), epithelial and immune cell type density showed variation across the three predefined clusters (Figure 5D and 5E). Although the small number of samples prevented formal statistical inference in relation to cell type density stack plots the results were overall in keeping with the earlier mIF results. Cluster 1 had the greatest proportion of T and B cells (6% and 11% respectively) whilst the patient from Cluster 2 had the lowest (1.4% and 0.9%), although the relative proportions of T cell subtypes were similar across the clusters (Figure 5E). While the overall proportion of macrophages and monocytes was similar, nearest neighbour analysis suggests differences in co-localisation of T cells and macrophages between Cluster 1 and Cluster 3. Cluster 1 showed a modest positive peak at radius 0.02-0.03, followed by predominantly negative values, indicating predominant spatial separation between CD68^+^ macrophages and CD8^+^ T cells. In contrast, in Cluster 3 there was a pronounced short-range spatial association between CD68^+^ macrophages and CD8^+^ T cells, with a strong positive clustering signal peak at r=0.1(Figure 5F). Given these results, gene expression amongst CD8^+^, CD4^+^ T cells and CD68^+^ macrophages were compared between Cluster 1 and Cluster 3 (Figure 5G).

There was little differential gene expression when CD8^+^ T cells were compared between Cluster 1 and Cluster 3, however CD4^+^ T cells in Cluster 3 overexpressed genes associated with antigen presentation and MHC II activity (HLA-DPB1, HLA-DRA, CD74). Macrophages in Cluster 3 relatively over-expressed genes including APOC1, APOE, A2M, CTSD, DUSP2, GPX1, LGMN and RNASE1, while those in Cluster 1 relatively over-expressed genes including ADAM8, CXCL8, CTSL, IL2RA, LMNA, MCL1, MMP12, SLC11A1, TFRC, TGFBI and TYMP. Although there was differential expression, almost all of these genes have been described to induce an immune-suppressive or pro-tumour microenvironment when expressed by TAMs in established solid cancers [16].

## Discussion

Colonic polyps, of which conventional adenomas are the most common, are a key risk factor for future polyps and colorectal cancer, necessitating surveillance in a proportion of those affected, and colonoscopic polypectomy for prevention [17]. However, despite stratification in terms of future risk, and evidence that surveillance is effective [18], patients with high-risk lesions remain at higher risk of future CRC even in modern cohorts [19].

This study reports that immune infiltrates in adenomas within a large bowel screening cohort of 2642 patients were associated with the development of metachronous polyps and colorectal cancer within a 6 year surveillance period. The initial counterintuitive finding that high CD3^+^ T cell density was associated with higher metachronous lesion risk (Figure 2) led to novel adenoma-immune clusters derived from mIF data which suggested three phenotypes; 1) high T cell density, low macrophage density with lowest risk, 2) immune excluded with highest risk, and 3) an intermediate risk group with high T cell, macrophage and αSMA density (Figure 4). These groups were not significantly associated with any previously described patient, exposure, or adenoma intrinsic risk factors [20], but were associated with future risk. Single cell spatially resolved analysis of gene expression across the immune infiltrate further revealed that the macrophages within the lamina propria generally expressed genes associated with an immune suppressive and pro-tumour microenvironment (Figure 5). However, the distance between these CD68^+^ macrophages and T cells within the adenoma lamina propria differed between adenomas from patients in Clusters 1 where there was spatial separation, and Cluster 3 where they were co-located to a greater degree. This suggests that innate-adaptive immune interactions within adenomas may not only be associated with lesion progression, but also future lesion risk even after removal.

Conventional adenomas are thought to arise from Wnt driven expansion of canonical stem cells, characterised by LGR5 expression and found at the crypt base, brought about by a well described sequence of mutations in genes, including *APC*, *KRAS* and *SMAD4,* with subsequent *p53* mutation leading to cancer characterised by chromosomal instability [21]. However, this process is incompletely understood, and lesion progression is dependent on other factors, including the local immune response [22]. Indeed, the presence of an immune exclusion phenotype within adenomas has been described as key to progression from pre-cancer to invasive disease [7]. In the present study there was an association between adenoma epithelial nuclear B catenin protein staining intensity using IHC and immune infiltrate, with the highest B catenin H-scores found in the immune excluded high metachronous risk group. Furthermore, a recent study in a large cohort of mixed histology colonic polyps showed that conventional adenomas with more advanced features, including villous morphology, had a lower epithelial CD8^+^ T cell density than those with less advanced features [9]. Although individual lesion progression, and metachronous lesion risk are different clinical outcomes, they may be linked by common fundamental processes like failure of effective immunoediting [23].

Adenomas removed by polypectomy at colonoscopy do not confer further direct risk to the patient. However, it may be that a wider functional immune field change or dysregulation effect exists [24]. Indeed, in a small subset of patients within this study, there was a modest relationship between CD3+ T cell density in the index and then metachronous lesion. If there is to be any consideration given to using immune-specific targets for prevention, then evidence of such dysregulation will have to be found in synchronous adenomas at colonoscopy, and indeed in the background apparently normal colonic mucosa [25]. Whilst it may be that immune dysregulation is a primary event, it may also be driven by the patient’s exposome, microbiome or underlying polygenic risk [26]. Understanding the immunological link which underpins these and future adenoma and cancer risk may be key to providing effective risk stratification and colorectal cancer prevention.

This study has a number of limitations. Serrated lesions were excluded as they are much less common than conventional adenomas, and their BRAF, RAS and CpG island methylation-driven biology generates a very different immune infiltrate to that of conventional adenomas [27]. The study is retrospective and despite the inclusion of exposure data and correlated clinical and pathological risk variables, no causal relationship can be drawn between the immune infiltrate observations and metachronous lesion outcome. All patients were from a single health board and external validation of these findings is required.

Furthermore, although NHS GGC is the largest such board in the UK, the majority of patients are Caucasian, therefore, the findings may not be generalisable in other countries and ethnic groups. Although the measured CD3^+^ T cell density in FFPE adenoma whole slide sections and TMA sections is correlated, there could be measurement error introduced by the need for serial sectioning. Similarly, the use of mIF may introduce further measurement error when compared to singleplex IHC. The number of samples analysed using CosMx was small, preventing useful comparison between the best (Cluster 1) and worst (Cluster 2) clinical outcome groups in terms of single cell spatial transcriptomic data. Finally, it must be considered that the use of mIF, RNAseq and spatial transcriptomic technologies would not be currently feasible in the clinical setting, however, the potential to generate simplified IHC or H&E-based classifiers for immune infiltrate phenotypes may allow such assessment in established bowel screening and surveillance programmes [28].

In conclusion, this study demonstrates that immune architecture within adenomas is associated with risk of metachronous polyp or colorectal cancer. These findings highlight immune contexture as a potential biomarker for colorectal surveillance and target for pre-cancer interception. As interest in precision screening, risk stratification for surveillance and the manipulation of immune responses in the pre-cancer interception space increases, understanding these mechanisms will be essential to translate discovery into clinical practice.

## Data availability statement

Anonymised clinical and pathological data are available on the secure Glasgow Safe Haven TRE platform. Access can be arranged by application to the authors and via the Glasgow SafeHaven TRE following ethical approval and the completion of mandatory information governance modules. Mutational and transcriptomic are available publicly at (E-MTAB-15349, E-MTAB-15346) and IHC data access can be discussed on reasonable application to the corresponding author.

## Ethics

Ethical approval was obtained for data (GSH/20/CO/002) and tissue analysis (22/WS/0020).

## Supporting information

Bulk transcriptomics code

CosMx code

## Acknowledgements

The authors thank Glasgow Tissue Research Facility for constructing the adenoma TMAs in this work, specifically Dr Pamela McCall, Dr Jennifer Hay, Dr Hannah Morgan and Mr Scott Murray. We thank Greater Glasgow and Clyde Biorepository including Ms Clare Orange for help in identifying and retrieving archival pathology tissue. We thank Mr William Sloan of NHS Greater Glasgow and Clyde for help in patient identification. We thank Prof John LeQuesne’s lab team at the University of Glasgow for assistance in scanning mIF slides.

## Contributors

STM, GL and JE conceived and designed the study. TI, AA, SSFA, LI, CKD, MD and CS performed the experiments, STM, TI, AA, JE, SSFA, LI, AL, GRM, MD, CS and NF analysed and interpreted the results. STM supervised the statistical analysis. NM provided expert pathological input and specimen review. STM wrote the manuscript, and TI, AA, SSFA, PD, NM, NBJ, MJ, GL, and JE critically reviewed the manuscript. STM, GL and JE supervised the study. GL provided administrative and project management support. ECP managed the PPIE aspects. All authors critically read and approved the paper. STM is the guarantor of the manuscript.

## Funding

Innovate UK: Awards 10054829 and 105858

Guts UK: ECR2023_03

Medical Research Scotland: PhD Studentship - PhD-50246-2020

Chief Scientist Office: Early Postdoctoral Fellowship - EPD/25/14

Cancer Research UK Scotland Institute: CTRQQR-2021\100006

## Patient and public involvement

The protocol and results of this study were presented to the Glasgow Colorectal PPIE group (https://www.gla.ac.uk/research/az/incise/forpatientsandthepublic/). The authors thank the group members for their contribution to the wording and emphasis of the reporting and their ongoing help with dissemination.

## Pre-print publication

A pre-print of this article was published on medRxiv on 02/03/2026

## MEDRXIV/2026/346992

https://medrxiv.org/cgi/content/short/2026.02.25.26346992v1

---

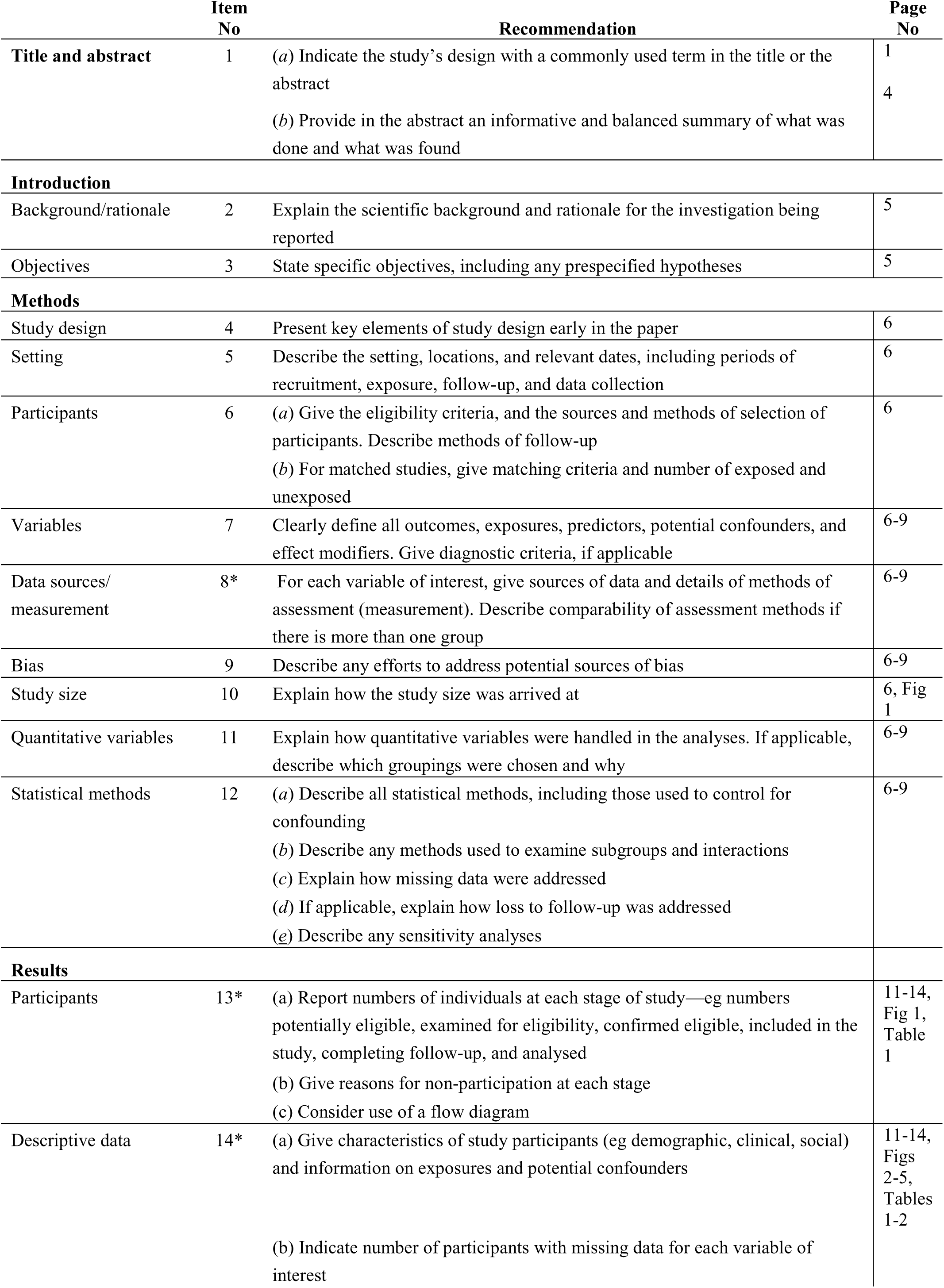
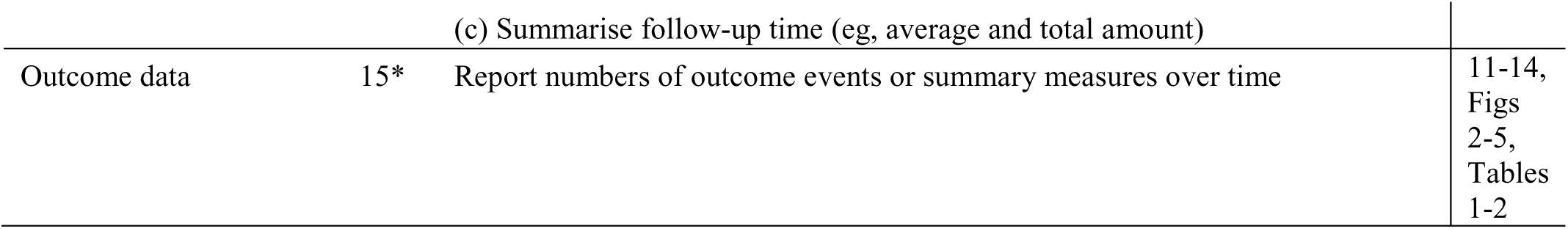
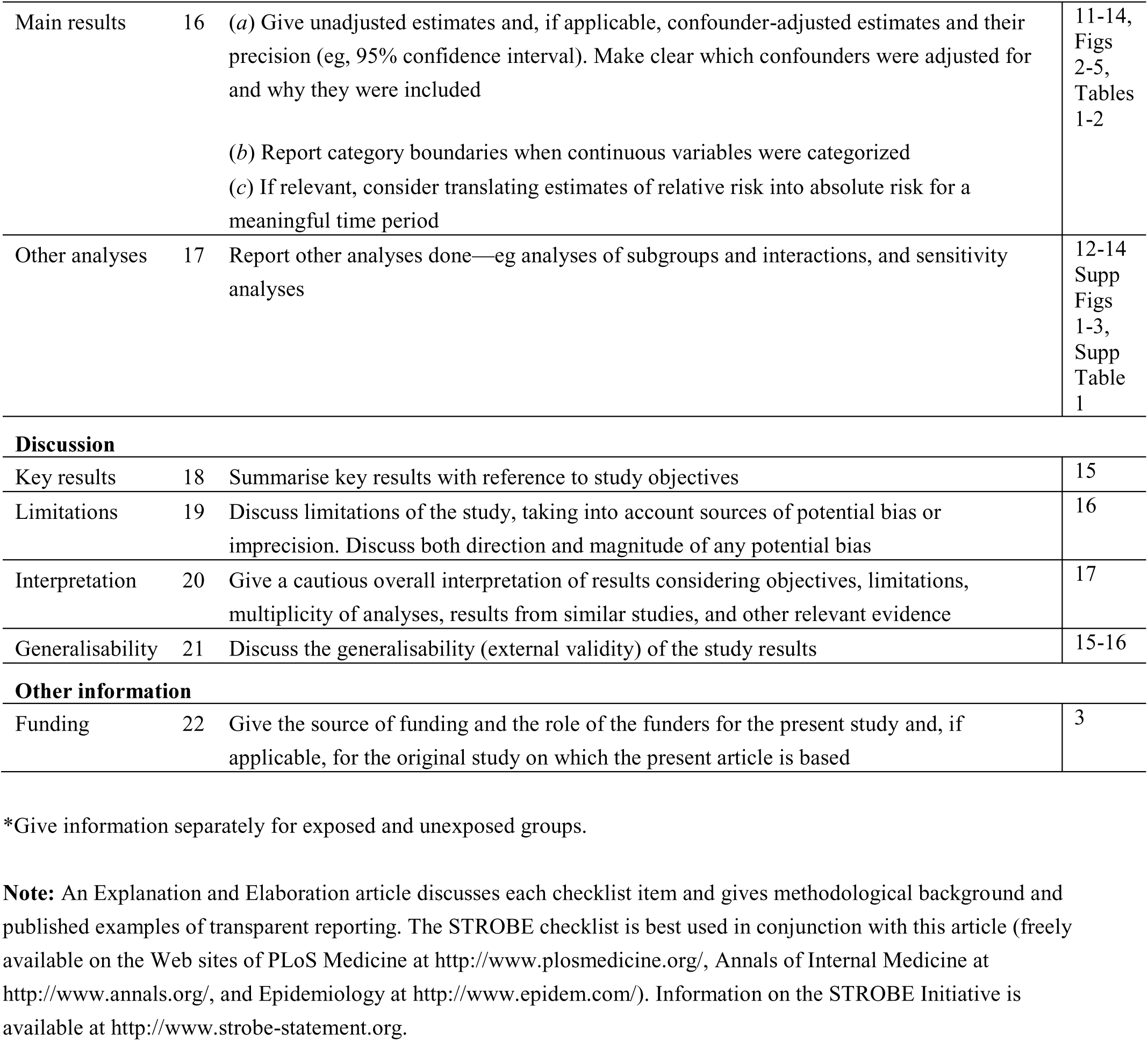
STROBE Statement—Checklist of items that should be included in reports of *cohort studies*.

