## Supplementary material for "Novel adenoma-immune phenotypes are associated with risk of metachronous polyps and colorectal cancer in a bowel screening cohort": Bulk transcriptomics code

```
# McSorley et al Study - INCISE Script ----

## Date: 2026-02-24
## Code (Originator): Natalie C. Fisher
## Code (Moderator): Sudhir B. Malla

## Description: The code for INCISE Bulk dataset, which includes initial QC, batch correction and normalisation, as well as data analysis such as MCPcounter for the study.

## ===========================================================================================

## 0. Load Library ----
library(tidyverse)
library(janitor)
library(dplyr)
library(tibble)
library(M3C)
library(sva)
library(edgeR)
library(ggpubr)
library(WGCNA)
library(preprocessCore)
library(MCPcounter)


## 1. Import data ----
folder1 <- file.path("C:/Users/3057008/Dropbox/02_Postdoc/projects/project_polyp2025/03_shared/Natalie/INCISE/Analysis/Final workflow single matrix/Preprocessing/Filter probes")
data <- read.delim(file = file.path(folder1, "INCISE_TempOSeq_raw_all.txt"), sep=",")
head(data)[1:5]


## 2. Filtering Probes ----

## find number of probes with 0 exp across all
data_t <- column_to_rownames(data, var="Study_ID")
## transpose so that genes are rows
data_t <- as.data.frame(t(data_t))
PROBES <- rownames(data_t)


data_t$sums <- rowSums(data_t)
range(data_t$sums) ## 0 276850744

## genes with 0 counts
sums_zero <- data_t %>% filter(data_t$sums == 0)
rownames(sums_zero)


## get the data again
data_t <- column_to_rownames(data, var="Study_ID")
data_t <- as.data.frame(t(data_t))


## get measure of variance of probe acrsoss all samples
variance <- sapply(data[c(2:22538)], var)
range(variance) ## 0 3864368595
variance <- as.data.frame(variance)
summary(variance)
 #    variance
 # Min.   :0.000e+00
 # 1st Qu.:6.400e+01
 # Median :9.650e+02
 # Mean   :4.913e+05
 # 3rd Qu.:1.211e+04
 # Max.   :3.864e+09

## genes with 0 variance
no_var <- variance %>% filter(variance == 0)


## using bottom 25th quartile to filter out variance
bottom_quartile <- variance %>% filter(variance < 6.400e+01)
nrow(bottom_quartile) ## 5640

top_3_quartiles <- variance %>% filter(variance > 6.400e+01)
nrow(top_3_quartiles) ## 16897


## extarct out samples for each matrix
data_2 <- column_to_rownames(data, var = "Study_ID")

## bottom_quartile
bottom_quartile_IDs <- rownames(bottom_quartile)
bottom_quartile_data <- data_2[, bottom_quartile_IDs]


## top_3_quartiles
top_3_quartiles_IDs <- rownames(top_3_quartiles)
top_3_quartiles_data <- data_2[, top_3_quartiles_IDs]


## transpose each
bottom_quartile_data_t <- as.data.frame(t(bottom_quartile_data))
top_3_quartiles_data_t <- as.data.frame(t(top_3_quartiles_data))
top_3_quartiles_data_t_2 <- rownames_to_column(top_3_quartiles_data_t, var="Probe_ID")

## save
# write.table(top_3_quartiles_data_t_2, "INCISE_raw_all_samples_topquantile_probevar.txt", sep=",", quote = F, row.names = F)


## 3. Filtering samples ----

## import data
data <- top_3_quartiles_data_t_2
data <- column_to_rownames(data, var="Probe_ID")


## histogrma of total reads
total_reads <- colSums(data)
hist(total_reads, breaks = 1000)

total_reads_df <- as.data.frame(total_reads)
## plot
p1 <- ggplot(total_reads_df, aes(total_reads)) +
  geom_histogram( color="black", binwidth = 5000) +
  geom_vline(xintercept = 2283865, linetype=3, color="red", size=1) +
  labs( title = "", x="Library size", y="Count") +
  theme(panel.background=element_blank(),
        panel.grid.major=element_line(),
        panel.grid.minor=element_line(),
        panel.border=element_rect(fill = NA),
        plot.title = element_text(hjust = 0.5, size = 14, face = 'bold'),
        axis.title.y=element_text(size = 16),
        axis.text.y = element_text(size = 16, colour = "black"),
        axis.title.x = element_text(size = 16),
        axis.text.x = element_text(size = 16, colour = "black"),
        legend.title = element_text(size = 14, face = 'bold', hjust = 0.7),
        legend.background = element_blank(),
        legend.key = element_rect(fill = 'white'),
        legend.text = element_text(size = 14))
print(p1)


## summary of total reads
summary(total_reads)
#  Min.  1st Qu.   Median     Mean  3rd Qu.     Max.
# 21164  4442502  5195942  5130785  5953782 12589515


## find 2 SD from mean of total counts
sd_val <- sd(total_reads) ## 1423460
mean_val <- mean(total_reads) ## 5130785

## cut off at 2 SD
sd_cutoff <- sd_val * 2  ## 2846920

## mean - 2SD
mean_sdcutoff_diff <- mean_val - sd_cutoff ## 2283865


## filter samples and see how it looks
total_read_df <- as.data.frame(total_reads)
data_t <- as.data.frame(t(data))
data_total_reads <- merge.data.frame(data_t, total_read_df, by=0)

data_total_reads_low_fitlered <- data_total_reads %>% filter(total_reads > mean_sdcutoff_diff)
nrow(data_total_reads_low_fitlered) ## 2532; 110 samples removed

hist(data_total_reads_low_fitlered$total_reads, breaks = 1000)

## plot
p1 <- ggplot(data_total_reads_low_fitlered, aes(total_reads)) +
  geom_histogram( color="black", binwidth = 5000) +
  labs( title = "", x="Library size", y="Count") +
  scale_x_continuous(limits = c(0, 14000000)) +
  theme(panel.background=element_blank(),
        panel.grid.major=element_line(),
        panel.grid.minor=element_line(),
        panel.border=element_rect(fill = NA),
        plot.title = element_text(hjust = 0.5, size = 14, face = 'bold'),
        axis.title.y=element_text(size = 16),
        axis.text.y = element_text(size = 16, colour = "black"),
        axis.title.x = element_text(size = 16),
        axis.text.x = element_text(size = 16, colour = "black"),
        legend.title = element_text(size = 14, face = 'bold', hjust = 0.7),
        legend.background = element_blank(),
        legend.key = element_rect(fill = 'white'),
        legend.text = element_text(size = 14))
print(p1)


## look at IDs and rank based on the total reads
data_total_reads_low_fitlered_2 <- data_total_reads_low_fitlered[c(1, 16899)]
data_total_reads_low_fitlered_2 <- data_total_reads_low_fitlered_2 [order(data_total_reads_low_fitlered_2$total_reads),]
data_total_reads_low_fitlered_2$rank <- 1:2532
## save
# write.table(data_total_reads_low_fitlered_2 , "INCISE_total_read_count_low_samples.txt", quote = F, row.names = F, sep=",")

## final matrix - removing the total reads column
data_total_reads_low_fitlered_export <- data_total_reads_low_fitlered[c(1:16898)]
colnames(data_total_reads_low_fitlered_export)[1] <- "Sample_ID"
## save
# write.table(data_total_reads_low_fitlered_export , "INCISE_raw_all_samples_3quartiles_probevar_low_samples.txt", quote = F, row.names = F, sep=",")


## 4. CombatSeq - Batch correction ----


## import data
data <- data_total_reads_low_fitlered_export
data <- column_to_rownames(data, var="Sample_ID")

## batch info
folder2 <- file.path("C:/Users/3057008/Dropbox/02_Postdoc/projects/project_polyp2025/03_shared/Natalie/INCISE/Analysis/Final workflow single matrix/Preprocessing/Combat Seq")
batch <- read.delim(file = file.path(folder2, "INCISE_batch_Bioclavis_Aula.txt"), sep=",")


## Extract out batches
IDs <- rownames(data)
nrow(data) ## 2532

## batch ids that matches the data
batch_data <- batch[batch$Sample_ID %in% IDs,]
nrow(batch_data) ## 2529


## one ID is missing
setdiff(IDs, batch$Sample_ID)
## "INC0684" "INC0698" "INC1689"


## extract out data for cases witha matched batch
batch_data_IDs <- batch_data$Sample_ID
length(batch_data_IDs) ## 2529
data_2 <- data[batch_data_IDs,]

## check in same order
all(rownames(data_2) == batch_data$Sample_ID)

table(batch_data$lib)


## remove lib0518 batches
lib0518 <- batch_data %>% filter(lib == "lib0518")
ID_remove <- lib0518$Sample_ID
data_3 <- data_2 %>% filter(!rownames(data_2) %in% ID_remove)

batch_data_2 <- batch_data %>% filter(lib != "lib0518")

data_3_t <- as.data.frame(t(data_3))

## quick pca
pca(data_3_t, labels = batch_data_2$lib, dotsize = 2)


## Run combat seq
batch_vec <- batch_data_2$lib

set.seed(321)
data_combat_seq <- ComBat_seq(counts = as.matrix(data_3_t), batch = batch_vec)
# Found 16 batches
# Using null model in ComBat-seq.
# Adjusting for 0 covariate(s) or covariate level(s)
# Estimating dispersions
# Fitting the GLM model
# Shrinkage off - using GLM estimates for parameters
# Adjusting the data


pca(data_combat_seq, labels = batch_data_2$lib, dotsize = 2)


## save
# write.table(data_combat_seq, "INCISE_raw_all_samples_3quartiles_probevar_low_samples_combatseq.txt", sep=",")


## 5. Probes-to-gene collapse (WGCNA) ----

data <- as.data.frame(data_combat_seq)
data[1:5, 1:5]

data <- rownames_to_column(data, var="Probe_ID")


## Gene symbol - remove strings from probes
data$gene_symbol <- gsub("_.*", "", data$Probe_ID)
data <- data[c(2529, 1:2528)]


## for duplicates look at the correlation of the probes for each one from batches code
## duplicates
data_dup <- data[duplicated(data$gene_symbol),]
nrow(data_dup) ## 1904 values duplicated

## unique genes
length(unique(data$gene_symbol)) ## 14993

## how many times each gene occur and identify any above > 1
n_occur <- data.frame(table(data$gene_symbol))
Dup_IDs <- n_occur[n_occur$Freq > 1,]
nrow(Dup_IDs) ## 1536
## get all the data for the duplicated genes
Dup_IDs_data <- data[data$gene_symbol %in% n_occur$Var1[n_occur$Freq > 1],]
nrow(Dup_IDs_data) ## 3440
## number of duplicates for each gene (i.e., 1 gene occurs 10 times)
table(Dup_IDs$Freq)
#    2    3    4    5    6    8   10
# 1251  225   46   11    1    1    1


## median more correlated when only 2 probes ##
## how many genes > 2 probes
more_then_2 <- Dup_IDs %>% filter(Dup_IDs$Freq > 2)
nrow(more_then_2) ## 285 genes
two_probes <- Dup_IDs %>% filter(Dup_IDs$Freq == 2) ## 1251 genes


## Collapse data using MaxMean function to remove of duplicated genes
rowGroup <- as.vector(data$gene_symbol)
rowID <- as.vector(data$Probe_ID)
rownames(data) <- NULL  ## remove rownames

datET <- column_to_rownames(data, var = "Probe_ID")[-1]

## run maxMean collapseRows
collapse.object <- collapseRows(datET=datET, rowGroup=rowGroup, rowID=rowID, method = "MaxMean")
Collapsed_genes <- data.frame(collapse.object$group2row, collapse.object$datETcollapsed)

head(Collapsed_genes)[1:5]
dim(Collapsed_genes) ## 14993  2529
Collapsed_genes <- Collapsed_genes[c(1, 3:2529)]

colnames(Collapsed_genes)[1] <- "Gene_symbol"


## save
# write.table(Collapsed_genes, "INCISE_raw_all_samples_3quartiles_probevar_low_samples_combatseq_maxMean.txt", quote=F, sep=",", row.names = F)


## 6. Quantile Normalisation and Log2+1 transformation ----

data <- as.data.frame(Collapsed_genes)
head(data)[1:5]

data[,1] <- NULL
data_matrix <- as.matrix(data)


## quantile normalisation function
quantile_normalisation <- function(df){
  df_rank <- apply(df,2,rank,ties.method="min")
  df_sorted <- data.frame(apply(df, 2, sort))
  df_mean <- apply(df_sorted, 1, mean)

  index_to_mean <- function(my_index, my_mean){
    return(my_mean[my_index])
  }

  df_final <- apply(df_rank, 2, index_to_mean, my_mean=df_mean)
  rownames(df_final) <- rownames(df)
  return(df_final)
}

## run quantile normalisation
QN_data <- quantile_normalisation(data_matrix)


## Log transformation (adding 1 pseudocount)
QN_log <- log2(QN_data +1)

range(QN_log) ## 0.00000 16.79459
range(QN_data) ## 0.0 113676.5


## into dataframe
QN_log <- as.data.frame(QN_log)
head(QN_log)[1:5]
QN_log <- rownames_to_column(QN_log, var="Gene_Symbol")


## save
# write.table(QN_log,"INCISE_raw_all_samples_3quartiles_probevar_low_samples_combatseq_maxMean_QN_all_M3_7_5.txt", sep=",", quote = F, row.names = F)


## 7. Data Analysis ----


### 7.1 MCPCounter ----

data <- QN_log
head(data)[1:5]
data <- column_to_rownames(data, var="Gene_Symbol")


## Run MCP
MCP_data <- MCPcounter.estimate(data,
                                featuresType="HUGO_symbols",
                                genes=read.table(curl("http://raw.githubusercontent.com/ebecht/MCPcounter/master/Signatures/genes.txt"),sep="\t",
                                                 stringsAsFactors=FALSE,header=TRUE,colClasses="character",check.names=FALSE))

MCP_data_df <- as.data.frame(MCP_data)
head(MCP_data_df)[1:5]


## save
# write.table(MCP_data_df, file="MCP_INCISE_entire_not_scaled.txt", quote=T, sep=",")


## Session End ----

## ===========================================================================================
```
