## Supplementary material for "Novel adenoma-immune phenotypes are associated with risk of metachronous polyps and colorectal cancer in a bowel screening cohort": CosMx code

```
# McSorley et al Study - INCISE Script ----

## Date: 2026-02-13
## Code (Originator): Assya Legrini
## Code (Moderator): Assya Legrini

## Description: The code for INCISE CosMx dataset, which includes initial QC, batch correction and normalisation, as well as data analysis

## ===========================================================================================
```

\# Libraries ----

library(Seurat)

library(ggplot2)

library(sctransform)

library(dplyr)

library(data.table)

library(tidyverse)

library(glmGamPoi)

library(utils)

library(Matrix)

library(InSituType)

library(DelayedMatrixStats)

library(stringr)

library(SpatialDecon)

library(scales)

library(CellPoly)

library(R.utils)

library(data.table)

library(pheatmap)

library(spatialTIME)

library(spatstat.geom)

library(decoupleR)

library(smiDE)

\# Load Qc'ed data ----

\## QC code in separate script

INCISE <- readRDS("E:/CosMx/INCISE_Seurat/INCISE-Lenient_QCed_seuratObject.RDS")

\# Normalization and Batch correction ------

set.seed(1234)

\## Normalize -----

INCISE <- NormalizeData(INCISE)

INCISE <- FindVariableFeatures(INCISE, selection.method = "vst", nfeatures = 2000)

INCISE <- ScaleData(INCISE)

gc()

\# Dimensional reduction

INCISE <- RunPCA(INCISE, assay = "RNA", reduction.name = "Unintegrated", features = VariableFeatures(INCISE))

elbow <- ElbowPlot(INCISE, ndims = 50, reduction = "Unintegrated")

elbow

INCISE <- RunUMAP(INCISE, reduction.name = "UnitegratedUMAP", reduction = "Unintegrated",dims = 1:30, verbose = FALSE)

gc()

\## Batch Correction ----

\### harmony integration -----

\# Integrate - using harmony

INCISE <- RunHarmony(INCISE,

assay.use = "RNA",

group.by.vars = c("ID"),

theta = 2, # diversity clustering penalty parameter - the larger the value, the more diverse the clusters, 2 is the default

reduction = "Unintegrated",

reduction.save = "Harmony")

\# Cluster after integration

INCISE <- FindNeighbors(INCISE, reduction = "Harmony",

dims = 1:30)

INCISE <- FindClusters(INCISE, reduction = "Harmony",

resolution = 0.5)

INCISE <- RunUMAP(INCISE, dims = 1:30,

reduction = "Harmony", reduction.name ="Integrated_UMAP")

\# save integrated and normalized data

metadata_cell <- read.csv("~/External_spatial_analysis/CosMx/INCISE/INCISE/INCISE_current_metadata.csv", row.names = 1)

colnames(metadata_cell)

metadata <- INCISE@meta.data

colnames(metadata)

metadata_cell <- metadata_cell %>% select(cell_id, panCK_gate, Semisup, Semisup_PanCkneg, mIF_cluster)

metadata <- metadata %>% left_join(metadata_cell, by = "cell_id")

INCISE <- AddMetaData(INCISE, metadata)

#saveRDS(INCISE, "E:/CosMx/INCISE_Seurat/INCISE_Seurat_Harmony-norm.RDS")

INCISE <- readRDS("D:/CosMx/External/INCISE_TMA_06_03_2025_11_50_26_565/INCISE_Seurat/INCISE_Seurat_Harmony-norm.RDS")

unique(INCISE@meta.data$mIF_cluster)

setwd("C:/Users/2426138/Documents/External_spatial_analysis/CosMx/INCISE/INCISE/Results/Final_figure")

Idents(INCISE) <- "Run_Tissue_name"

class(INCISE$mIF_cluster)

INCISE@meta.data$mIF_cluster <- as.character(INCISE@meta.data$mIF_cluster)

rename_map <- c(

"1" = "Cluster 1",

"2" = "Cluster 2",

"3" = "Cluster 3"

)

\# Apply the renaming

INCISE$mIF_cluster <- plyr::revalue(INCISE$mIF_cluster, replace = rename_map)

table(INCISE@meta.data$mIF_cluster)

\## Visualisation - UMAPs -----

pdf("INCISE_Harmonyintergration_UMAPs.pdf", width = 8, height = 8)

DimPlot(INCISE, reduction = "Unintegrated")

DimPlot(INCISE, reduction = "Integrated_UMAP")

DimPlot(INCISE, reduction = "UnitegratedUMAP", group.by = "ID")

DimPlot(INCISE, reduction = "Integrated_UMAP", group.by = "ID")

DimPlot(INCISE, reduction = "Unintegrated", group.by = "mIF_cluster")

DimPlot(INCISE, reduction = "Integrated_UMAP", group.by = "mIF_cluster")

\# axis features removed

p <- DimPlot(INCISE, reduction = "Integrated_UMAP", group.by = "ID", pt.size = 1.5)

p + theme(

axis.text = element_blank(),

axis.ticks = element_blank()

#axis.title = element_blank()

)

p <- DimPlot(INCISE, reduction = "Integrated_UMAP", group.by = "mIF_cluster", pt.size = 1.5)

p + theme(

axis.text = element_blank(),

axis.ticks = element_blank(),

#axis.title = element_blank()

)

dev.off()

\### png version ----

tiff("UMAP_unintegrated_ID_UMAP.tif")

p <- DimPlot(INCISE, reduction = "UnitegratedUMAP", group.by = "ID", pt.size = 1.5)

p + theme(

axis.text = element_blank(),

axis.ticks = element_blank()

#axis.title = element_blank()

)

dev.off()

tiff("Harmony_ID_UMAP.tif")

p <- DimPlot(INCISE, reduction = "Integrated_UMAP", group.by = "ID", pt.size = 1.5)

p + theme(

axis.text = element_blank(),

axis.ticks = element_blank()

#axis.title = element_blank()

) + NoLegend()

dev.off()

tiff("Harmony_ID_UMAP_legend.tif")

p <- DimPlot(INCISE, reduction = "Integrated_UMAP", group.by = "ID", pt.size = 1.5)

p + theme(

axis.text = element_blank(),

axis.ticks = element_blank()

#axis.title = element_blank()

)

dev.off()

tiff("Harmony_mIF_cluster_UMAP.tif")

p <- DimPlot(INCISE, reduction = "Integrated_UMAP", group.by = "mIF_cluster", pt.size = 1.5)

p + theme(

axis.text = element_blank(),

axis.ticks = element_blank()

#axis.title = element_blank()

) + NoLegend()

dev.off()

tiff("Harmony_mIF_cluster_UMAP_legend.tif")

p <- DimPlot(INCISE, reduction = "Integrated_UMAP", group.by = "mIF_cluster", pt.size = 1.5)

p + theme(

axis.text = element_blank(),

axis.ticks = element_blank()

#axis.title = element_blank()

)

dev.off()

\# Celltyping IO panel CosMx ----

\# Semi-supervised clustering using IO panel

counts <- INCISE@assays$RNA$counts %>%

t() %>% as("dgCMatrix")

counts[1:5, 1:5]

Negative <- as.matrix(INCISE@assays[["negprobes"]]@counts) %>%

t()

Negative[1:5, 1:5]

negmean <- Matrix::rowMeans(Negative)

\# extrat out UMAP from seurat object

INCISE_umap <- INCISE@reductions[['Integrated_UMAP']]@cell.embeddings

rm(INCISE)

\# Profiles ----

\# IO CosMx panel

IO <- read.csv("~/Nanostring GeoMx/CosMx/Cell_typing/CosMx_IO_profiles.csv", row.names=1)

IO %>% as.matrix()

IO[1:5, 1:5]

\# this bit is needed to match the colours by the cell profile names

cols <- c('#B3DE69', '#FCCDE5', '#D9D9D9', '#BC80BD', '#CCEBC5',

'#FFED6F', '#E41A1C', '#377EB8', '#4DAF4A', '#984EA3',

'#FF7F00', '#FFFF33', '#A65628', '#F781BF', '#999999',

'#4B0082', '#4682B4', '#D2B48C', '#008080', '#D8BFD8',

'#FF6347', '#40E0D0', '#EE82EE', '#F5DEB3', '#FFFFFF',

'#A52A2A', '#DEB887', '#5F9EA0', '#7FFF00', '#D2691E',

'#FF4500', '#2E8B57', '#8B0000', '#483D8B', '#2F4F4F'

)

\# Create a named vector to map colors to column names

IO_cols_1K <- setNames(cols[1:ncol(IO)], colnames(IO))

print(IO_cols_1K)

\# WTA

data(ioprofiles)

str(ioprofiles)

ioprofiles[1:5, 1:10]

data("iocolors")

print(iocolors)

gc()

\# Anchor stats -----

\## pick the preferred profile

astats <- get_anchor_stats(counts = counts,

neg = negmean,

profiles = IO)

#saveRDS(astats, file = "anchor_stats_INCISE-TMA.rds", compress = "xz")

anchor <- readRDS("anchor_stats_INCISE-TMA.rds")

negmean.per.totcount <- mean(rowMeans(Negative)) / mean(rowSums(counts))

per.cell.bg <- rowSums(counts) * negmean.per.totcount

\## Anchors ----

anchors <- choose_anchors_from_stats(counts = counts,

neg = negmean,

bg = per.cell.bg,

anchorstats = astats,

n_cells = 10000,

min_cosine = 0.4,

min_scaled_llr = 0.03,

insufficient_anchors_thresh = 5)

check <- anchors %>% as.data.frame()

unique(check$.)

table(check$.)

\# anchors on UMAP

par(mfrow = c(1, 1))

plot(INCISE_umap, pch = 16, cex = 0.1, col = "peachpuff1", xaxt = "n", yaxt = "n", xlab = "", ylab = "",

main = "Semi-supervised anchor cells")

points(INCISE_umap[!is.na(anchors), ], col = IO_cols_1K[anchors[!is.na(anchors)]], pch = 16, cex = 0.6)

legend("topright", pch = 16, col = IO_cols_1K[setdiff(unique(anchors), NA)],

legend = setdiff(unique(anchors), NA), cex = 0.65)

updatedprofiles <- updateReferenceProfiles(reference_profiles = IO,

counts = counts,

neg = negmean,

bg = per.cell.bg,

anchors = anchors)

\# Will actually gate for IF - just checking to see what the anchors are and how well immune cells are being picked up

immunofluordata <- matrix(rpois(n = nrow(counts) * 4, lambda = 100),

nrow(counts))

cohort <- fastCohorting(immunofluordata,

gaussian_transform = TRUE)

\# Semi-sup ------

semisup <- insitutype(

x = counts,

neg = negmean,

cohort = cohort,

bg = NULL,

n_clusts = 5:41,

reference_profiles = updatedprofiles$updated_profiles,

update_reference_profiles = FALSE,

n_phase1 = 10000,

n_phase2 = 20000,

n_phase3 = 1e+05,

n_starts = 1,

max_iters = 40

)

\# save so that it doens't need to be repeated

wd <- "E:/CosMx/INCISE_Seurat/"

file_path <- file.path(wd, "INCISE_semisupervised_IO.RDS")

saveRDS(semisup, file = file_path)

semisup <- readRDS("D:/CosMx/External/INCISE_TMA_06_03_2025_11_50_26_565/INCISE_Seurat/INCISE_semisupervised_IO.RDS")

str(semisup)

head(semisup[["clust"]])

semisup$clust <- recode(

semisup$clust,

"a" = "Epithelium.A",

"b" = "Epithelium.B",

"c" = "Epithelium.C",

"d" = "Epithelium.D",

"e" = "Epithelium.E"

)

\# define cluster colors:

cols <- c('#B3DE69', '#FCCDE5', '#D9D9D9', '#BC80BD', '#CCEBC5',

'#FFED6F', '#E41A1C', '#377EB8', '#4DAF4A', '#984EA3',

'#FF7F00', '#FFFF33', '#A65628', '#F781BF', '#999999',

'#4B0082', '#4682B4', '#D2B48C', '#008080', '#D8BFD8',

'#FF6347', '#40E0D0', '#EE82EE', '#F5DEB3', '#FFFFFF',

'#A52A2A', '#DEB887', '#5F9EA0', '#7FFF00', '#D2691E',

'#FF4500', '#2E8B57', '#8B0000', '#483D8B', '#2F4F4F'

)

cols <- cols[seq_along(unique(semisup$clust))]

names(cols) <- unique(semisup$clust)

cols[is.element(names(cols), names(iocolors))] <- iocolors[names(cols)[is.element(names(cols), names(iocolors))]]

\## Flightpath ----

fp <- flightpath_plot(insitutype_result = semisup, col = cols[semisup$clust], showclusterconfidence = TRUE)

print(fp)

\# compute the flightpath layout:

fp_layout <- flightpath_layout(logliks = semisup$logliks, profiles = semisup$profiles)

#str(fp_layout)

\# plot it:

par(mar = c(0,0,0,0))

plot(fp_layout$cellpos, pch = 16, cex = 0.5, col = cols[semisup$clust], xaxt = "n", yaxt = "n", xlab = "", ylab = "",

main = "Flightpath Semi-supervised IO panel",)

text(fp_layout$clustpos, rownames(fp_layout$clustpos), cex = 0.5)

\## Visualisation - Celltype UMAPs and flight path -----

colnames(INCISE@meta.data)

cols <- c('#B3DE69', '#FCCDE5', '#D9D9D9', '#BC80BD', '#CCEBC5',

'#FFED6F', '#E41A1C', '#377EB8', '#4DAF4A', '#984EA3',

'#FF7F00', '#FFFF33', '#A65628', '#F781BF', '#999999',

'#4B0082', '#4682B4', '#D2B48C', '#008080', '#D8BFD8',

'#FF6347', '#40E0D0', '#EE82EE', '#F5DEB3', '#FFFFFF',

'#A52A2A', '#DEB887', '#5F9EA0', '#7FFF00', '#D2691E',

'#FF4500', '#2E8B57', '#8B0000', '#483D8B', '#2F4F4F'

)

celltype <- INCISE@meta.data$Semisup_final %>% unique()

cols <- setNames(cols[1:length(celltype)], celltype)

print(cols)

\### pdf version -----

pdf("Semisup_IO_panel_celltype_UMAP-flightpath.pdf", width = 10, height = 10)

p <- DimPlot(INCISE, reduction = "Integrated_UMAP", group.by = "Semisup_final", cols = cols, pt.size = 1.5,

label = TRUE, label.box = TRUE, repel = TRUE)

p + theme(

axis.text = element_blank(),

axis.ticks = element_blank()

#axis.title = element_blank()

)

p + theme(

axis.text = element_blank(),

axis.ticks = element_blank()

#axis.title = element_blank()

) + NoLegend()

fp <- flightpath_plot(insitutype_result = semisup, col = cols[semisup$clust], showclusterconfidence = TRUE)

print(fp)

dev.off()

\### tiff version ----

tiff("Semisup_IO_panel_celltype_UMAP.tif")

p + theme(

axis.text = element_blank(),

axis.ticks = element_blank()

#axis.title = element_blank()

) + NoLegend()

dev.off()

tiff("Semisup_IO_panel_celltype_UMAP_legend.tif")

p + theme(

axis.text = element_blank(),

axis.ticks = element_blank()

#axis.title = element_blank()

)

dev.off()

tiff("Semisup_flightpath.tif")

fp <- flightpath_plot(insitutype_result = semisup, col = cols[semisup$clust], showclusterconfidence = TRUE)

print(fp)

dev.off()

\## Visualisation - Density Barcharts per cell type -----

metadata <- INCISE@meta.data

colnames(metadata)

pdf("Barchart_densities.pdf")

\# all cell types

cell_cluster <- metadata %>% group_by(mIF_cluster, Semisup_final) %>%

summarise(Nb = n()) %>%

mutate(C = sum(Nb)) %>%

mutate(percent = Nb/C*100)

write.csv(cell_cluster, "All_celltype_density_protortions.csv")

ggplot(cell_cluster, aes(x = mIF_cluster, y = percent, fill = Semisup_final)) +

geom_bar(stat = "identity") +

scale_fill_manual(values = cols) +

labs(title = "Semisup final cell types", x = "mIF cluster", y = "Percentage") +

theme_minimal() +

theme(axis.text.x = element_text(hjust = 1))

\# filtered cell types

unique(metadata$Semisup_final)

mIF_celltypes <- c('Fibroblast', 'Fibroblast.aSMA', 'Monocyte', 'Macrophage','Macrophage.CD68',

'T.cell.CD4', 'T.cell.regulatory', 'T.cell.CD8')

metadata_filterd <- metadata %>% filter(Semisup_final %in% mIF_celltypes)

cell_cluster <- metadata_filterd %>% group_by(mIF_cluster, Semisup_final) %>%

summarise(Nb = n()) %>%

mutate(C = sum(Nb)) %>%

mutate(percent = Nb/C*100)

write.csv(cell_cluster, "Filtered_celltype_density_protortions.csv")

ggplot(cell_cluster, aes(x = mIF_cluster, y = percent, fill = Semisup_final)) +

geom_bar(stat = "identity") +

scale_fill_manual(values = cols) +

labs(title = "Semisup final cell types filtered - per mIF cluster", x = "mIF_cluster", y = "Percentage") +

theme_minimal() +

theme(axis.text.x = element_text(hjust = 1))

dev.off()

\### tiff version -----

tiff("Barchart_all_cells.tif")

cell_cluster <- metadata %>% group_by(mIF_cluster, Semisup_final) %>%

summarise(Nb = n()) %>%

mutate(C = sum(Nb)) %>%

mutate(percent = Nb/C*100)

ggplot(cell_cluster, aes(x = mIF_cluster, y = percent, fill = Semisup_final)) +

geom_bar(stat = "identity") +

scale_fill_manual(values = cols) +

labs(title = "Semisup final cell types", x = "mIF cluster", y = "Percentage") +

theme_minimal() +

theme(axis.text.x = element_text(hjust = 1))

dev.off()

tiff("Barchart_all_cells_NoLegend.tif")

cell_cluster <- metadata %>% group_by(mIF_cluster, Semisup_final) %>%

summarise(Nb = n()) %>%

mutate(C = sum(Nb)) %>%

mutate(percent = Nb/C*100)

ggplot(cell_cluster, aes(x = mIF_cluster, y = percent, fill = Semisup_final)) +

geom_bar(stat = "identity") +

scale_fill_manual(values = cols) +

labs(title = "Semisup final cell types", x = "mIF cluster", y = "Percentage") +

theme_minimal() +

theme(axis.text.x = element_text(hjust = 1)) +

NoLegend()

dev.off()

tiff("Barchart_filtered_cells.tif")

mIF_celltypes <- c('Fibroblast', 'Fibroblast.aSMA', 'Monocyte', 'Macrophage','Macrophage.CD68',

'T.cell.CD4', 'T.cell.regulatory', 'T.cell.CD8')

metadata_filterd <- metadata %>% filter(Semisup_final %in% mIF_celltypes)

cell_cluster <- metadata_filterd %>% group_by(mIF_cluster, Semisup_final) %>%

summarise(Nb = n()) %>%

mutate(C = sum(Nb)) %>%

mutate(percent = Nb/C*100)

ggplot(cell_cluster, aes(x = mIF_cluster, y = percent, fill = Semisup_final)) +

geom_bar(stat = "identity") +

scale_fill_manual(values = cols) +

labs(title = "Semisup final cell types filtered - per mIF cluster", x = "mIF_cluster", y = "Percentage") +

theme_minimal() +

theme(axis.text.x = element_text(hjust = 1))

dev.off()

tiff("Barchart_filtered_cells_NoLegend.tif")

mIF_celltypes <- c('Fibroblast', 'Fibroblast.aSMA', 'Monocyte', 'Macrophage','Macrophage.CD68',

'T.cell.CD4', 'T.cell.regulatory', 'T.cell.CD8')

metadata_filterd <- metadata %>% filter(Semisup_final %in% mIF_celltypes)

cell_cluster <- metadata_filterd %>% group_by(mIF_cluster, Semisup_final) %>%

summarise(Nb = n()) %>%

mutate(C = sum(Nb)) %>%

mutate(percent = Nb/C*100)

ggplot(cell_cluster, aes(x = mIF_cluster, y = percent, fill = Semisup_final)) +

geom_bar(stat = "identity") +

scale_fill_manual(values = cols) +

labs(title = "Semisup final cell types filtered - per mIF cluster", x = "mIF_cluster", y = "Percentage") +

theme_minimal() +

theme(axis.text.x = element_text(hjust = 1)) +

NoLegend()

dev.off()

\# mIF cluster specific UMAPs and example core -----

\# mIF clusters combined UMAP

DimPlot(INCISE, reduction = "Integrated_UMAP", group.by = "mIF_cluster")

\## cluster 1 UMAPs ----

set.seed(1234)

unique(metadata$mIF_cluster)

cells_to_keep <- rownames(INCISE@meta.data)[INCISE@meta.data$mIF_cluster == "Cluster 1"]

INCISE_mIF1 <- subset(INCISE, cells = cells_to_keep)

INCISE_mIF1 <- FindNeighbors(INCISE_mIF1, reduction = "Harmony", dims = 1:30)

INCISE_mIF1 <- FindClusters(INCISE_mIF1, reduction = "Harmony",

resolution = 0.5)

INCISE_mIF1 <- RunUMAP(INCISE_mIF1, dims = 1:30,

reduction = "Harmony", reduction.name ="IntegratedmIF1_UMAP")

\### Visualization ----

tiff("Semisup_IO_panel_celltype_UMAP_cluster1.tif")

p <- DimPlot(INCISE_mIF1, reduction = "IntegratedmIF1_UMAP", group.by = "Semisup_final", cols = cols, pt.size = 1.5,

label = TRUE, label.box = TRUE, repel = TRUE)

p + theme(

axis.text = element_blank(),

axis.ticks = element_blank()

#axis.title = element_blank()

) + NoLegend()

dev.off()

\## cluster 2 UMAPs -----

set.seed(1234)

cells_to_keep <- rownames(INCISE@meta.data)[INCISE@meta.data$mIF_cluster == "Cluster 2"]

INCISE_mIF2 <- subset(INCISE, cells = cells_to_keep)

INCISE_mIF2 <- FindNeighbors(INCISE_mIF2, reduction = "Harmony", dims = 1:30)

INCISE_mIF2 <- FindClusters(INCISE_mIF2, reduction = "Harmony",

resolution = 0.5)

INCISE_mIF2 <- RunUMAP(INCISE_mIF2, dims = 1:30,

reduction = "Harmony", reduction.name ="Integrated_mIF2_UMAP")

\### Vizualisation ----

tiff("Semisup_IO_panel_celltype_UMAP_cluster2.tif")

p <- DimPlot(INCISE_mIF2, reduction = "Integrated_mIF2_UMAP", group.by = "Semisup_final", cols = cols, pt.size = 1.5,

label = TRUE, label.box = TRUE, repel = TRUE)

p + theme(

axis.text = element_blank(),

axis.ticks = element_blank()

#axis.title = element_blank()

) + NoLegend()

dev.off()

\## cluster 3 UMAPs -----

set.seed(1234)

cells_to_keep <- rownames(INCISE@meta.data)[INCISE@meta.data$mIF_cluster == "Cluster 3"]

INCISE_mIF3 <- subset(INCISE, cells = cells_to_keep)

INCISE_mIF3 <- FindNeighbors(INCISE_mIF3, reduction = "Harmony", dims = 1:30)

INCISE_mIF3 <- FindClusters(INCISE_mIF3, reduction = "Harmony",

resolution = 0.5)

INCISE_mIF3 <- RunUMAP(INCISE_mIF3, dims = 1:30,

reduction = "Harmony", reduction.name ="Integrated_mIF3_UMAP")

\### Visualization ----

tiff("Semisup_IO_panel_celltype_UMAP_cluster3.tif")

p <- DimPlot(INCISE_mIF3, reduction = "Integrated_mIF3_UMAP", group.by = "Semisup_final", cols = cols, pt.size = 1.5,

label = TRUE, label.box = TRUE, repel = TRUE)

p + theme(

axis.text = element_blank(),

axis.ticks = element_blank()

#axis.title = element_blank()

) + NoLegend()

dev.off()

\# DEA -----

\## CD8 ----

INCISE_CD8 <- subset(INCISE, subset = Semisup_final %in% c("T.cell.CD8"))

INCISE_CD8 <- subset(INCISE_CD8, subset = mIF_cluster %in% c("1", "3"))

metadata <- INCISE_CD8@meta.data

Idents(object = INCISE_CD8) <- INCISE_CD8@meta.data$mIF_cluster

gc()

CD8_top_markers <-FindAllMarkers(INCISE_CD8, test.use = "MAST")

celltype <- "CD8"

CD8_top_markers$Celltype <- celltype

png(filename="DEA_mIFcluster1Vs3_CD8.png")

EnhancedVolcano(CD8_top_markers,

lab = "gene",

ylab = bquote(~-Log[10]~italic(P)),

x = 'avg_log2FC',

y = 'p_val_adj',

title = 'mIF clusters: 1 vs 3',

pCutoff = 0.05,

FCcutoff = 1,

pointSize = 3.0,

labSize = 3.0)

dev.off()

#write.csv(CD8_top_markers, "CD8_markers_DEA.csv")

\## CD4 -----

INCISE_CD8 <- subset(INCISE, subset = Semisup_final %in% c("T.cell.CD4"))

INCISE_CD8 <- subset(INCISE_CD8, subset = mIF_cluster %in% c("1", "3"))

metadata <- INCISE_CD8@meta.data

Idents(object = INCISE_CD8) <- INCISE_CD8@meta.data$mIF_cluster

gc()

CD4_top_markers <-FindAllMarkers(INCISE_CD8, test.use = "MAST")

celltype <- "CD4"

CD4_top_markers$Celltype <- celltype

png(filename="DEA_mIFcluster1Vs3_CD4.png")

EnhancedVolcano(CD4_top_markers,

lab = "gene",

ylab = bquote(~-Log[10]~italic(P)),

x = 'avg_log2FC',

y = 'p_val_adj',

title = 'mIF clusters: 1 vs 3',

pCutoff = 0.05,

FCcutoff = 1,

pointSize = 3.0,

labSize = 3.0)

dev.off()

#write.csv(CD4_top_markers, "CD4_markers_DEA.csv")

\## Tregs ----

INCISE_CD8 <- subset(INCISE, subset = Semisup_final %in% c("T.cell.regulatory"))

INCISE_CD8 <- subset(INCISE_CD8, subset = mIF_cluster %in% c("1", "3"))

metadata <- INCISE_CD8@meta.data

Idents(object = INCISE_CD8) <- INCISE_CD8@meta.data$mIF_cluster

gc()

foxp3_top_markers <-FindAllMarkers(INCISE_CD8, test.use = "MAST")

celltype <- "Tregs"

foxp3_top_markers$Celltype <- celltype

png(filename="DEA_mIFcluster1Vs3_Treg.png")

EnhancedVolcano(foxp3_top_markers,

lab = "gene",

ylab = bquote(~-Log[10]~italic(P)),

x = 'avg_log2FC',

y = 'p_val_adj',

title = 'mIF clusters: 1 vs 3',

pCutoff = 0.05,

FCcutoff = 1,

pointSize = 3.0,

labSize = 3.0)

dev.off()

#write.csv(foxp3_top_markers, "Tregs_markers_DEA.csv")

\## Macrophages ----

INCISE_CD8 <- subset(INCISE, subset = Semisup_final %in% c("Macrophage.CD68"))

INCISE_CD8 <- subset(INCISE_CD8, subset = mIF_cluster %in% c("1", "3"))

metadata <- INCISE_CD8@meta.data

Idents(object = INCISE_CD8) <- INCISE_CD8@meta.data$mIF_cluster

gc()

Mac_top_markers <-FindAllMarkers(INCISE_CD8, test.use = "MAST")

celltype <- "Macrophage"

Mac_top_markers$Celltype <- celltype

png(filename="DEA_mIFcluster1Vs3_Macrophages.png")

EnhancedVolcano(Mac_top_markers,

lab = "gene",

ylab = bquote(~-Log[10]~italic(P)),

x = 'avg_log2FC',

y = 'p_val_adj',

title = 'mIF clusters: 1 vs 3',

pCutoff = 0.05,

FCcutoff = 1,

pointSize = 3.0,

labSize = 3.0)

dev.off()

#write.csv(Mac_top_markers, "Macrophage_markers_DEA.csv")

\## fibroblasts -----

INCISE_CD8 <- subset(INCISE, subset = Semisup_final %in% c("Fibroblast.aSMA"))

INCISE_CD8 <- subset(INCISE_CD8, subset = mIF_cluster %in% c("1", "3"))

metadata <- INCISE_CD8@meta.data

Idents(object = INCISE_CD8) <- INCISE_CD8@meta.data$mIF_cluster

gc()

fib_top_markers <-FindAllMarkers(INCISE_CD8, test.use = "MAST")

celltype <- "Fibroblast"

fib_top_markers$Celltype <- celltype

png(filename="DEA_mIFcluster1Vs3_Fibrolast.png")

EnhancedVolcano(fib_top_markers,

lab = "gene",

ylab = bquote(~-Log[10]~italic(P)),

x = 'avg_log2FC',

y = 'p_val_adj',

title = 'mIF clusters: 1 vs 3',

label

pCutoff = 0.05,

FCcutoff = 1,

pointSize = 3.0,

labSize = 3.0)

dev.off()

#write.csv(fib_top_markers, "Firboblast_markers_DEA.csv")

\# Nearest neighbour analysis ----

\## Ran on HPC

metadata <- INCISE@meta.data

unique(metadata$ID)

metadata$x_slide_mm <- as.numeric(metadata$x_slide_mm )

metadata$y_slide_mm <- as.numeric(metadata$y_slide_mm )

metadata$unique_fov <- paste0("Patient_", metadata$ID)

\# create false classifier roughly by cell types - immune and non-immune

unique(metadata$Semisup_final)

metadata <- metadata %>%

mutate(

Classifier.Label = case_when(

Semisup_final %in% c("a", "b", "c", "d", "e") ~ "Epithelium",

TRUE ~ "TME"

)

)

#extract and make long data frame of relevant spatial information

spatial_data <- metadata %>%

select( unique_fov, ID,

x_slide_mm, y_slide_mm, #cell location

Classifier.Label, Semisup_final) %>% #cell cluster

mutate(positive = 1) %>% #add column for positivity of the cells

arrange(Semisup_final) %>% #arrange cells clusters in alphabetical order

pivot_wider(names_from = "Semisup_final",

values_from = "positive",

values_fill = 0)

#convert to spatial list

metadata %>%

group_by(ID) %>%

summarise(

x_min = min(x_slide_mm),

x_max = max(x_slide_mm),

y_min = min(y_slide_mm),

y_max = max(y_slide_mm)

)

spatial_list <- split(spatial_data, spatial_data$unique_fov)

#create sample summary level data

cell_types <- unique(metadata$Semisup_final)

summary_df <- lapply(spatial_list, function(spat){

#need to collapse to the number of positive of each cell type and

#percent of cells positive for each cell type

spat %>%

group_by(unique_fov ,ID) %>% #can remove cell locations here for sample level information

summarise(total_cells = n(),

across(!!cell_types, sum)) %>%

mutate(across(!!cell_types, ~ .x /total_cells * 100, .names = "percent_{col}")) #percent of total

}) %>%

do.call(bind_rows, .)

#create the clinical data

#this is also sample level data

colnames(metadata)

clinical <- metadata %>%

select(unique_fov, ID, cell_id, mIF_cluster, slide_ID_numeric, assay_type, Run_Tissue_name, Classifier.Label,

Panel, version, contains("qc"), x_slide_mm, y_slide_mm, median_negprobes:percOfDataFromErrorPerCell) %>%

distinct() %>%

mutate(patient_id = ID, .before = 1)

clinical$x_slide_mm <- as.numeric(clinical$x_slide_mm )

clinical$y_slide_mm <- as.numeric(clinical$y_slide_mm )

\## SpatialTIME - Build Object ----

#create mif

spat_obj = create_mif(clinical_data = clinical,

sample_data = summary_df,

spatial_list = spatial_list,

patient_id = "ID",

sample_id = "unique_fov")

\# Check that all spatial list entries have an 'ID' column

all(sapply(spatial_list, function(df) "unique_fov" %in% names(df)))

\# Check coordinate ranges again (just to be sure)

range(metadata$x_slide_mm)

range(metadata$y_slide_mm)

#saveRDS(spat_obj, "mif_spatial_object_INCISE.rds")

spat_obj

#change source location

table(metadata$Semisup_final)

\# NaN is present in the derived counted - needs to be removed

unique(metadata$Semisup_final)

names(spatial_list[[1]])

lapply(spatial_list, function(df) {

colSums(df[, c("T.cell.CD8", "Macrophage.CD68",

"T.cell.CD4", "T.cell.regulatory")])

})

\# 2mm cores

spat_obj3 <- bi_NN_G(mif = spat_obj,

mnames = c('T.cell.CD8', 'Macrophage.CD68',

'T.cell.CD4', 'Macrophage.CD68',

'T.cell.regulatory', 'Macrophage.CD68'), # list of phenotypes to compare for mIF clusters

num_permutations = 500,

edge_correction = 'rs',

r = seq(0, 0.5, by = 0.001), # take into account the radius want to minimise severe edge effect in TMA cores

workers = 1,

overwrite = TRUE,

xloc = "x_slide_mm", yloc = "y_slide_mm")

\# see where G function plateaus

plot(spat_obj3$derived$bivariate_NN$r,

spat_obj3$derived$bivariate_NN$`Observed G`)

#saveRDS(spat_obj3, "Bivariate_nearest-neighbour_tcells-macrophages_new.rds")

#spat_obj3 <- readRDS("Bivariate_nearest-neighbour_tcells-macrophages.rds")

replace_nan_with_na <- function(x) {

if (is.data.frame(x) || is.matrix(x)) {

x[] <- lapply(x, function(col) ifelse(is.nan(col), NA, col))

} else if (is.numeric(x)) {

x[is.nan(x)] <- NA

}

return(x)

}

\# Apply this function to each element of `spat_obj3$derived`

spat_obj3$derived <- map(spat_obj3$derived, replace_nan_with_na)

bivariate_NN <- spat_obj3$derived$bivariate_NN %>% na.omit()

clinical <- spat_obj3[["clinical"]] %>% select(ID, unique_fov, cell_id, mIF_cluster)

range(bivariate_NN$`Degree of Clustering Permutation`)

colnames(bivariate_NN)

gc()

\# add mIF clusters

mIF_cluster <- clinical %>% select(unique_fov, ID, mIF_cluster)

\# there is an NA cluster - change this to zero

mIF_cluster[is.na(mIF_cluster)] <- 0

mIF_cluster_unique <- mIF_cluster %>% distinct(unique_fov, .keep_all = TRUE)

bivariate_NN <- bivariate_NN %>% left_join(mIF_cluster_unique, by = "unique_fov")

mIF_cluster <- unique(bivariate_NN$mIF_cluster)

color_palette <- RColorBrewer::brewer.pal(n = length(mIF_cluster), name = "Set3")

names(color_palette) <- mIF_cluster

\## Visualization Bivariate Nearest ----

pdf("Spatial_analysis_Bi-Nearest-neighbour_Tcell-macrophages_mIF-clusters.pdf")

\### unique_FOV ----

bivariate_NN <- bivariate_NN %>%

filter(mIF_cluster != "0") %>%

mutate(mIF_cluster = as.factor(mIF_cluster))

\# Unique Counted markers

counted_markers <- unique(bivariate_NN$Counted)

\# colour palette - matching umap

colour_palette <- c("1" = "#F8766D",

"2" = "#4DAF4A",

"3" = "#377EB8")

\# Generate and store all plots in a named list

bi_nn_plots <- map(counted_markers, function(marker) {

bivariate_NN %>%

filter(Anchor == "Macrophage.CD68", Counted == marker) %>%

ggplot(aes(x = r, y = `Degree of Clustering Permutation`, color = mIF_cluster)) +

geom_line(show.legend = TRUE) +

theme_bw() +

labs(title = paste("From Macrophage.CD68 to", marker)) +

scale_color_manual(values = colour_palette)

}) %>%

set_names(counted_markers) # Name the list by marker

bi_nn_plots[["T.cell.CD8"]]

bi_nn_plots[["T.cell.CD4"]]

bi_nn_plots[["T.cell.regulatory"]]

\### Summary per cluster ----

bivariate_summary <- bivariate_NN %>%

group_by(mIF_cluster, Anchor, Counted, r) %>%

summarise(across(where(is.numeric), mean, na.rm = TRUE), .groups = "drop")

write.csv(bivariate_summary, "Average_Bivariate_nearest-neighbour_tcells-macrophages.csv")

bi_nn_plots <- map(counted_markers, function(marker) {

bivariate_summary %>%

filter(Anchor == "Macrophage.CD68", Counted == marker) %>%

ggplot(aes(x = r, y = `Degree of Clustering Permutation`, color = mIF_cluster)) +

geom_line(show.legend = TRUE) +

theme_bw() +

labs(title = paste("From Macrophage.CD68 to", marker)) +

scale_color_manual(values = colour_palette)

}) %>%

set_names(counted_markers) # Name the list by marker

bi_nn_plots[["T.cell.CD8"]]

bi_nn_plots[["T.cell.CD4"]]

bi_nn_plots[["T.cell.regulatory"]]

dev.off()

\# Cellpoly plots ----

\# Define the cell IDs to filter by

gc()

metadata <- read.csv("INCISE_current_metadata.csv")

patientID <- unique(metadata$ID)

INC0003 <- metadata %>% filter(ID == "INC0221")

filtered_cells <- INC0003$cell_id

filtered_cells_file <- "filtered_cells.txt"

\# Path to the compressed .csv.gz file

file_path <- "E:/CosMx/External/INCISE_TMA_06_03_2025_11_50_26_565/INCISE_TMA_FF/GRIINCTMAR6KJE21112024/GRIINCTMAR6KJE21112024_tx_file.csv.gz"

\# Create a function to filter rows during file read

filter_large_file <- function(file_path, filtered_cells, chunk_size = 1e5) {

\# Open the gzipped file

con <- gzfile(file_path, "r")

\# Read the header to get column names

header <- readLines(con, n = 1)

column_names <- strsplit(header, ",")[[1]]

\# Initialize an empty data.table for filtered results

result <- data.table::data.table(matrix(ncol = length(column_names), nrow = 0))

setnames(result, column_names)

\# Read the file in chunks

while (TRUE) {

\# Read the next chunk

lines <- readLines(con, n = chunk_size)

if (length(lines) == 0) break # Stop when no lines are left

\# Convert the chunk to a data.table

chunk <- fread(

text = paste(c(header, lines), collapse = "\n"),

header = TRUE, showProgress = FALSE

)

\# Filter the chunk by 'cell' column

filtered_chunk <- chunk[cell %in% filtered_cells]

\# Append filtered rows to the result

result <- rbind(result, filtered_chunk, fill = TRUE)

}

\# Close the file connection

close(con)

return(result)

}

\# Apply the function to filter the dataset

tx_filtered <- filter_large_file(file_path, filtered_cells)

#head(tx_filtered)

\# matching column names of cellpoly package

names(tx_filtered)[2] <- "cell"

names(tx_filtered)[3] <- "cell_id"

names(tx_filtered)[4] <- "x_local"

names(tx_filtered)[5] <- "y_local"

names(tx_filtered)[6] <- "x"

names(tx_filtered)[7] <- "y"

\## INC0003 ----

\## Cluster 1

\# filter transcript file for an example patient core - script found in INCISE folder called filtering test

INC0003_metadata <- metadata %>% filter(ID == "INC0003")

\# Generate colors for each unique Semisup_final type

cols <- c('#B3DE69', '#FCCDE5', '#D9D9D9', '#BC80BD', '#CCEBC5',

'#FFED6F', '#E41A1C', '#377EB8', '#4DAF4A', '#984EA3',

'#FF7F00', '#FFFF33', '#A65628', '#F781BF', '#999999',

'#4B0082', '#4682B4', '#D2B48C', '#008080', '#D8BFD8',

'#FF6347', '#40E0D0', '#EE82EE', '#F5DEB3', '#FFFFFF',

'#A52A2A', '#DEB887', '#5F9EA0', '#7FFF00', '#D2691E',

'#FF4500', '#2E8B57', '#8B0000', '#483D8B', '#2F4F4F'

)

cols <- cols[seq_along(unique(metadata$Semisup_final))]

names(cols) <- unique(metadata$Semisup_final)

cellcols <- cols

celltypes <- INC0003_metadata %>%

dplyr::select(cell_id, Semisup_final) %>%

deframe()

custom_data <- list(

transcript_df = tx_filtered,

celltype = celltypes,

cellcols = cellcols

)

frame()

legend("center",

pch = 16,

col = custom_data$cellcols,

legend = names(custom_data$cellcols),

cex = 0.4)

transcript_df <- custom_data$transcript_df

head(transcript_df)

\# creating polygon data

polys <- initPolys(cell_ids = unique(transcript_df$cell_id))

#polys[1:3]

polys <- cellPolys(polys = polys,

transcript_df = transcript_df,

type = "chull",

cell_ids = NULL)

#polys[1]

#names(polys)

\## Draw polygons ----

tiff("INC0003_polygon-core_celltypes.tif")

frame()

legend("center",

pch = 16,

col = custom_data$cellcols,

legend = names(custom_data$cellcols),

cex = 0.4)

drawPolys(polys,

cell_ids = NULL, #tempids,

alpha = 0.1,

col = scales::alpha(custom_data$cellcols[custom_data$celltype[names(polys)]], 0.5),

border = custom_data$cellcols[custom_data$celltype[names(polys)]],

add = FALSE, outputpolys = FALSE, xaxt = "n", yaxt = "n", xlab = "", ylab = "", asp = 1)

dev.off()

\## INC0010 ----

\## CLuster 2

\# filter transcript file for an example patient core - script found in INCISE folder called filtering test

INC0010_metadata <- metadata %>% filter(ID == "INC0010")

\# Generate colors for each unique Semisup_final type

cols <- c('#B3DE69', '#FCCDE5', '#D9D9D9', '#BC80BD', '#CCEBC5',

'#FFED6F', '#E41A1C', '#377EB8', '#4DAF4A', '#984EA3',

'#FF7F00', '#FFFF33', '#A65628', '#F781BF', '#999999',

'#4B0082', '#4682B4', '#D2B48C', '#008080', '#D8BFD8',

'#FF6347', '#40E0D0', '#EE82EE', '#F5DEB3', '#FFFFFF',

'#A52A2A', '#DEB887', '#5F9EA0', '#7FFF00', '#D2691E',

'#FF4500', '#2E8B57', '#8B0000', '#483D8B', '#2F4F4F'

)

cols <- cols[seq_along(unique(metadata$Semisup_final))]

names(cols) <- unique(metadata$Semisup_final)

cellcols <- cols

celltypes <- INC0010_metadata %>%

dplyr::select(cell_id, Semisup_final) %>%

deframe()

custom_data <- list(

transcript_df = tx_filtered,

celltype = celltypes,

cellcols = cellcols

)

frame()

legend("center",

pch = 16,

col = custom_data$cellcols,

legend = names(custom_data$cellcols),

cex = 0.4)

transcript_df <- custom_data$transcript_df

head(transcript_df)

\# creating polygon data

polys <- initPolys(cell_ids = unique(transcript_df$cell_id))

#polys[1:3]

polys <- cellPolys(polys = polys,

transcript_df = transcript_df,

type = "chull",

cell_ids = NULL)

#polys[1]

#names(polys)

\## Draw polygons ----

tiff("INC0010_polygon-core_celltypes.tif")

frame()

legend("center",

pch = 16,

col = custom_data$cellcols,

legend = names(custom_data$cellcols),

cex = 0.4)

drawPolys(polys,

cell_ids = NULL, #tempids,

alpha = 0.1,

col = scales::alpha(custom_data$cellcols[custom_data$celltype[names(polys)]], 0.5),

border = custom_data$cellcols[custom_data$celltype[names(polys)]],

add = FALSE, outputpolys = FALSE, xaxt = "n", yaxt = "n", xlab = "", ylab = "", asp = 1)

dev.off()

\## INC0221 ----

\## Cluster 3

\# filter transcript file for an example patient core - script found in INCISE folder called filtering test

INC0221_metadata <- metadata %>% filter(ID == "INC0221")

\# Generate colors for each unique Semisup_final type

cols <- c('#B3DE69', '#FCCDE5', '#D9D9D9', '#BC80BD', '#CCEBC5',

'#FFED6F', '#E41A1C', '#377EB8', '#4DAF4A', '#984EA3',

'#FF7F00', '#FFFF33', '#A65628', '#F781BF', '#999999',

'#4B0082', '#4682B4', '#D2B48C', '#008080', '#D8BFD8',

'#FF6347', '#40E0D0', '#EE82EE', '#F5DEB3', '#FFFFFF',

'#A52A2A', '#DEB887', '#5F9EA0', '#7FFF00', '#D2691E',

'#FF4500', '#2E8B57', '#8B0000', '#483D8B', '#2F4F4F'

)

cols <- cols[seq_along(unique(metadata$Semisup_final))]

names(cols) <- unique(metadata$Semisup_final)

cellcols <- cols

celltypes <- INC0221_metadata %>%

dplyr::select(cell_id, Semisup_final) %>%

deframe()

custom_data <- list(

transcript_df = tx_filtered,

celltype = celltypes,

cellcols = cellcols

)

frame()

legend("center",

pch = 16,

col = custom_data$cellcols,

legend = names(custom_data$cellcols),

cex = 0.4)

transcript_df <- custom_data$transcript_df

head(transcript_df)

\# creating polygon data

polys <- initPolys(cell_ids = unique(transcript_df$cell_id))

#polys[1:3]

polys <- cellPolys(polys = polys,

transcript_df = transcript_df,

type = "chull",

cell_ids = NULL)

#polys[1]

#names(polys)

\## Draw polygons ----

tiff("INC0221_polygon-core_celltypes.tif")

frame()

legend("center",

pch = 16,

col = custom_data$cellcols,

legend = names(custom_data$cellcols),

cex = 0.4)

drawPolys(polys,

cell_ids = NULL, #tempids,

alpha = 0.1,

col = scales::alpha(custom_data$cellcols[custom_data$celltype[names(polys)]], 0.5),

border = custom_data$cellcols[custom_data$celltype[names(polys)]],

add = FALSE, outputpolys = FALSE, xaxt = "n", yaxt = "n", xlab = "", ylab = "", asp = 1)

dev.off()
