## Supplementary tables and figures for "Novel adenoma-immune phenotypes are associated with risk of metachronous polyps and colorectal cancer in a bowel screening cohort"

**Supplementary Data**

**Supplementary tables:**

Supplementary Table 1: Association of Multiplex Immunofluorescence-based Adenoma Immune Clusters and Demographic, Clinical, and Pathological Characteristics in Bowel Screening Programme Polypectomy Patients

|  | | **Cluster 1**  **n (%)** | **Cluster 2**  **n (%)** | **Cluster 3**  **n (%)** | ***p*** |
| --- | --- | --- | --- | --- | --- |
| Sex | Male | 74 (30) | 82 (34) | 87 (36) | 0.134 |
|  | Female | 17 (19) | 34 (39) | 37 (42) |  |
| Age | Screening Age | 89 (98) | 112 (97) | 116 (93) | 0.271 |
| (years) | Above Screening Age | 2 (2) | 4 (3) | 8 (7) |  |
| BMI | <25 | 18 (32) | 21 (37) | 18 (32) | 0.634 |
| (kg/m^2^) | 25-30 | 29 (24) | 42 (35) | 49 (41) |  |
|  | >30 | 33 (30) | 41 (37) | 36 (33) |  |
| Smoking | Never | 33 (30) | 36 (32) | 43 (38) | 0.834 |
|  | Ex | 35 (25) | 51 (36) | 54 (39) | 0.834 |
|  | Current | 23 (30) | 29 (37) | 26 (33) |  |
| SIMD | 1 | 30 (30) | 37 (37) | 33 (33) | 0.474 |
| (quintile) | 2 | 14 (20) | 31 (45) | 24 (35) |  |
|  | 3 | 15 (33) | 15 (33) | 16 (34) |  |
|  | 4 | 12 (30) | 12 (30) | 16 (40) |  |
|  | 5 | 20 (27) | 21 (28) | 34 (45) |  |
| Aspirin | No | 79 (26) | 108 (36) | 112 (38) | 0.299 |
|  | Yes | 12 (39) | 8 (26) | 11 (36) |  |
| Statin | No | 76 (27) | 96 (34) | 107 (38) | 0.631 |
|  | Yes | 15 (29) | 20 (39) | 16 (31) |  |
| Adenoma | Right Colon | 4 (17) | 6 (25) | 14 (58) | 0.088 |
| Site | Left Colon | 81 (29) | 101 (37) | 94 (34) |  |
|  | Rectum | 5 (18) | 9 (32) | 14 (50) |  |
| Dysplasia | LGD | 77 (28) | 101 (37) | 99 (36) | 0.305 |
|  | HGD | 14 (26) | 15 (28) | 25 (46) |  |
| Histology | Tubular | 45 (35) | 45 (35) | 40 (30) | 0.044 |
|  | Tubulovillous | 44 (24) | 66 (36) | 73 (40) |  |
|  | Villous | 2 (11) | 5 (28) | 11 (61) |  |

Screening Age: 50-74 years. Above Screening Age: 75+ years. BMI: Body Mass Index. SIMD: Scottish Index of Multiple Deprivation. HGD: High-grade Dysplasia. LGD: Low-grade Dysplasia.

**Supplementary Figures:**

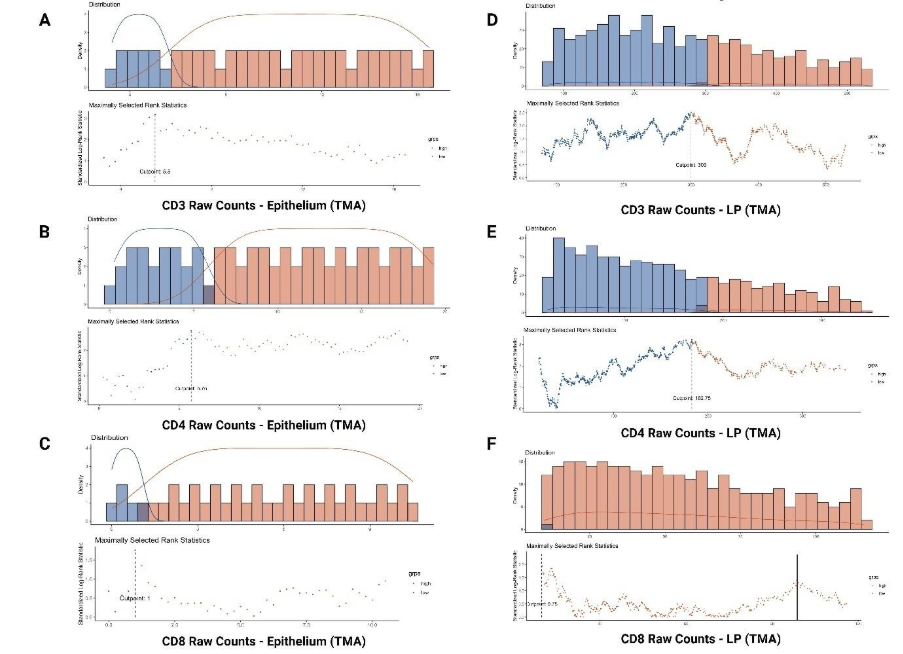

**Supplementary Figure 1**. Thresholding for Cell Density Dichotomisation. Raw cell density counts for epithelial (**A**) CD3, (**B**) CD4, and (**C**) CD8, and for (**D**) CD3, (**E**) CD4, and (**F**) CD8 in the lamina propria were cut off as low or high based on thresholds generated by *survminer* package in R. As CD8 in the lamina propria (F) had a very low threshold, the next highest peak was used as the cut-off point, marked by solid block line.

**A**

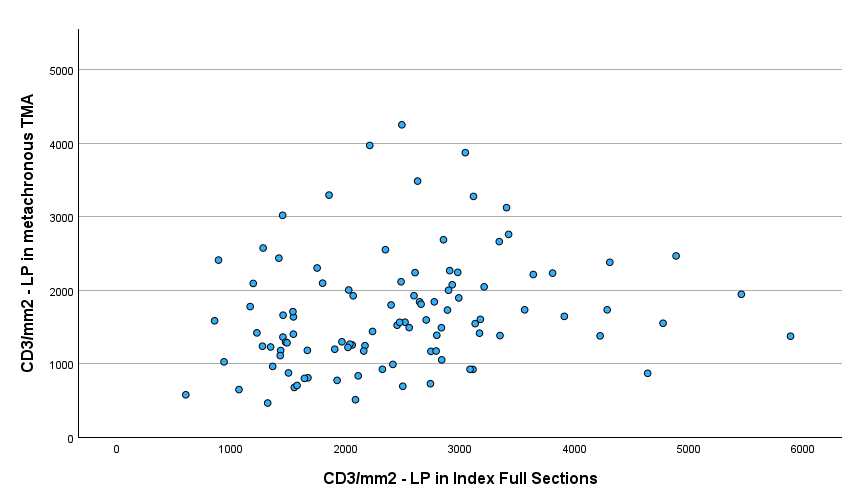

**B**

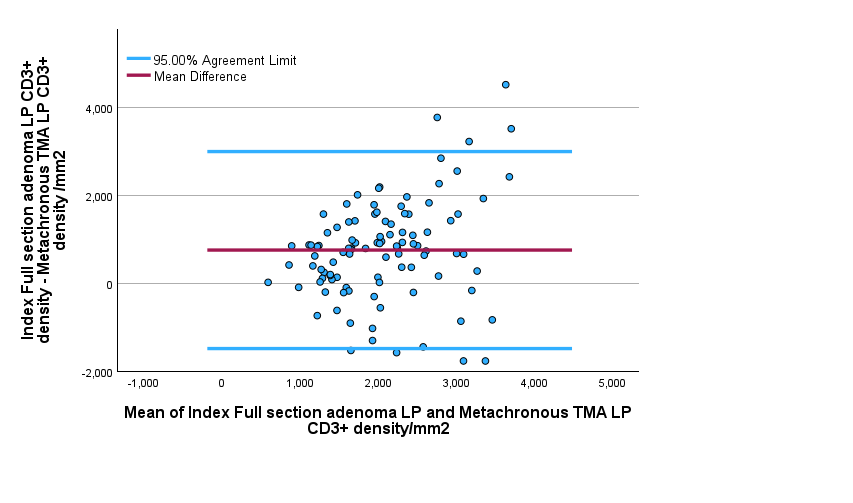

**Supplementary Figure 2**. (**A**) Scatter plot comparison of CD3+ cell density counts between index full section adenoma lamina propria and metachronous TMA adenoma lamina propria (n=102), showing statistically significant weak to moderate positive correlation (Spearman's rho=0.303, p=0.002). (**B**) Bland-Altman plot reporting mean difference and 95% limits of agreement between CD3+ cell density counts in index full section adenoma lamina propria and metachronous TMA adenoma lamina propria.

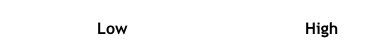

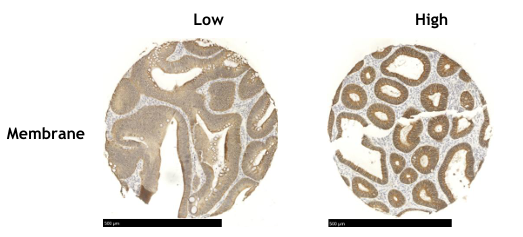

**B**

**A**

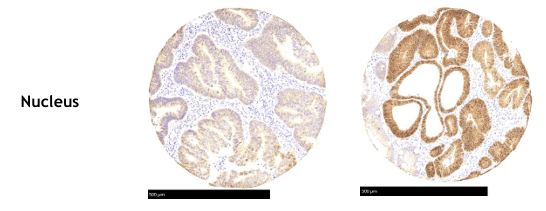

**D**
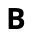

**C**
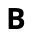

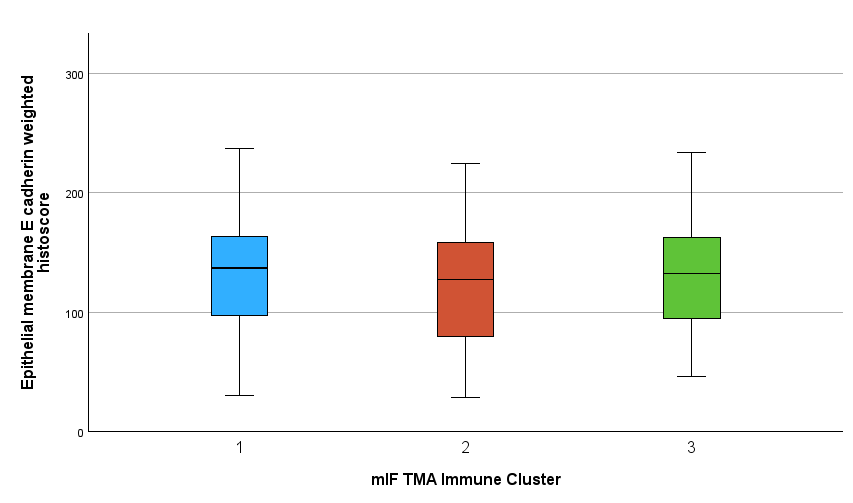

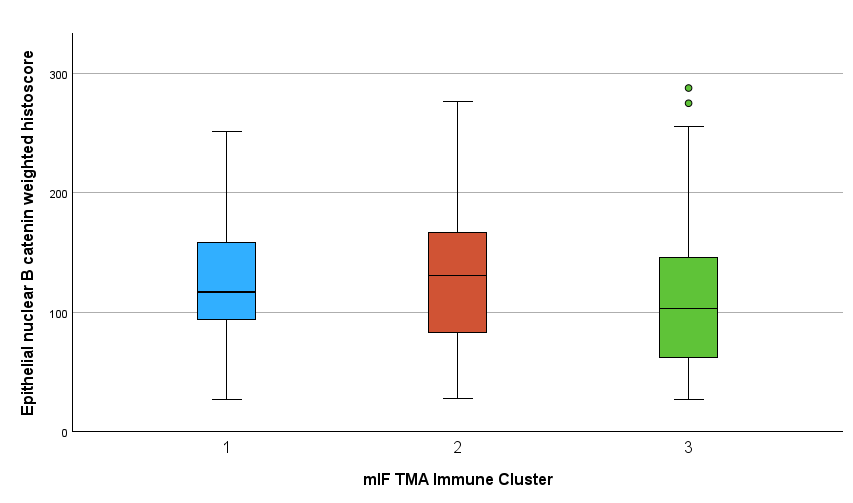

**Supplementary Figure 3**. Comparison of immunohistochemistry (IHC) nuclear B-Catenin and membranous E-cadherin staining in index adenoma TMA core epithelial cells, grouped by multiplex immunofluorescence (mIF) defined TMA immune clusters.  Examples of low and high staining intensity in 0.6mm TMA cores for (**A**) nuclear B catenin expression and (**B**) membranous E-cadherin expression. (**C**) Boxplot showing significantly different distribution (Kruskal Wallis p=0.006) of weighted histoscores of nuclear B catenin expression intensity across TMA cores (n=288) grouped into mIF immune cluster 1 (median H-score =117), cluster 2 (median H score =131), and cluster 3 (median H score =103). (**D**) Boxplot showing no significant difference in distribution (Kruskal Wallis p=0.507) of weighted histoscores of membranous E-cadherin expression intensity across TMA cores (n=299) grouped into mIF immune clusters 1 to 3.
