## Supplementary Methods for "Novel adenoma-immune phenotypes are associated with risk of metachronous polyps and colorectal cancer in a bowel screening cohort"

Patient cohort:

The INCISE cohort consists of 2642 bowel screening patients from NHS Greater Glasgow and Clyde (2009-2016) who on index colonoscopy had a minimum of one polyp removed and received at least one surveillance colonoscopy 6 months - 6 years after, with the primary outcome the presence or absence of a metachronous polyp(s) or CRC during that period (Johnstone *et al.* 2023). Recorded data included patient demography (age, sex, Scottish Index of Multiple Deprivation), prescribing data, co-morbidities, some exposures, endoscopy report results and histopathological data. These included number of polyps present at index and follow-up colonoscopies, polyp histology, morphology, location, size, and grade of dysplasia. From these variables the British Society of Gastroenterology (BSG) 2020 guideline risk categories were determined and recorded for each patient. Data were collected from electronic health records (Clinical Portal, Orion Health, Boston USA; Trakcare, Intersystem, Boston USA), endoscopy reporting software (Unisoft GI Reporting Software, v2.5, Unisoft Medical Systems, UK) and Telepath Laboratory Information Management System electronic pathology database. Patients with index serrated lineage lesions, CRC, history of CRC, diagnosed polyposis or CRC predisposition syndromes, inflammatory bowel disease, diagnosed autoimmune disease or systemic vasculitis were excluded.

Patient Tissue:

Patient tissue was requested from the NHS Greater Glasgow and Clyde Tissue Biorepository and cut by the Glasgow Tissue Research Facility (GTRF). Full sections (n=2466) were cut at a thickness of 4 μm, and TMAs were sectioned at a thickness of 2.5 μm. After sectioning tissue, slides were baked at 37℃ and stored at 4℃ until used. TMAs slides were baked for 1 hour at 60℃ before staining to reduce core loss during the staining process. TMAs were constructed using a TMA Grandmaster (3DHistech, Budapest, Hungary) to allow high throughput analysis of FFPE adenoma tissue while conserving usable sample. Each patient within a TMA was represented by up to four cores 0.6 mm in diameter and 2.5-µm thickness. A pathologist identified and annotated areas encompassing superficial luminal facing- and basal crypt epithelium within each adenoma. Two of 4 cores were taken from the luminal epithelium, and 2 from the basal epithelium to account for tissue heterogeneity.

Chromogenic single-plex Immunohistochemistry:

The PT Module (epredia, UK) with protocol was used to stain whole slides and TMAs with anti-CD3 (1:100), TMAs with anti-CD4 (NCL-L-CD4-368, Leica Biosystems, UK), and anti-CD8 (1:400, Clone CD/144B, Dako, Denmark), B-Catenin (1:1000, M3539, Dako, Denmark) and E-Cadherin (1:5000, 610182, BD Biosciences, USA) and Ultravision Quanto detection system HRP (Cat. TL-125-QHL, epredia, UK). TMAs were baked until wax melted, antigen retrieval was in Dewax and HIER buffer H (pH 9, Cat. TA-999-DHBH, epredia, UK) and washing in tris-buffered saline with tween (TBS-tween). Endogenous peroxidases were blocked for 10 min with Ultravision Hydrogen Peroxide Block (Cat. TA-125-H202Q, epredia, UK) and non-specific binding was blocked for 5 min with Ultravision Protein Block (Cat. TA-125-PBQ, epredia, UK). Slides were incubated for 30 min RT in CD3 diluted with Antibody Diluent OP Quanto (Cat. TA-125-ADQ, epredia, UK). TMAs were amplified with Primary Antibody Amplifier (Cat. TL-125-QPB, epredia, UK) for 10 min, after which Horse Radish Peroxidase (HRP) Polymer Quanto (Cat. TL-125-QPH, epredia, UK) was added for 10 min. 3,3′-Diaminobenzidine (DAB) Quanto (Cat. TA-125-QHDX, epredia, UK) was used for visualisation. Slides were counterstained with Haematoxylin Gill III (Cat. 3801540E, Leica Biosystems, USA), Acid Alcohol 0.5% (Cat. 3803650E, Leica Biosystems, USA), and Scott’s Tap Water Substitute (Cat. 3802901E, Leica Biosystems, USA). Slides were then dehydrated in increasing concentrations of alcohol (70%, 100%) and finally in analytical reagent and cleared in Histoclear II (Cat. HS-202, national diagnostics, USA), before being mounted and cover-slipped using ClearVue mounting medium (Epredia). All mounted slides were viewed under a brightfield microscope to assess staining quality and ensure the absence of staining artefacts and bubbles, which could negatively impact the scanning process. Any slides that failed this step had their coverslips gently removed (by submerging in xylene overnight at RT), placed in HistoClear-II and remounted. All mounted slides were cleaned of excess mounting medium using 100% ethanol and gently scraping off the excess using a 23G needle. Whole slides were scanned at x40 and TMA slides at x20 resolution in the Glasgow Tissue Research Facility (GTRF) with a Hamamatsu S60 Nanozoomer (Hamamatsu, Japan) and images were stored within NZConnect (v 1.1.0) and accessed using NDP Viewer.

Assessment of IHC:

Cell density (cells/mm^2^) was determined by point count of CD3^+^ cells in whole slide adenoma FFPE samples (n=2466) using Visiopharm apps (Visiopharm A/S, Hoersholm, Denmark) developed by OracleBio. The image analysis workflow was composed of four stages: (i) identification of viable tissue and major artefacts; (ii) tissue classification into adenoma and non-adenoma compartments; (iii) subdivision of adenoma/non-adenoma compartments into epithelium and lamina propria regions of interest (ROIs); and (iv) cell segmentation and classification (CD3^+^, CD3^-^) within each ROI (Figure 2C).

Quantification of T cells on TMA was performed using QuPath digital pathology software (Version 0.5.1) (Bankhead et al. 2017). TMA images were de-arrayed into a grid and cores with inadequate staining quality were excluded. Positive cell detection was then carried out to determine the number of CD3^+^, CD4^+^ and CD8^+^ cells in each core. Detection parameters were adjusted for detection image, pixel size, maximum cell size, sigma value. A Random Trees object classifier was trained using two annotations of polyp epithelium and lamina propria in a subset of cores, before applying to all other images. To ensure reliability up to 10% of the images were manually scored and intraclass correlation coefficients (ICCC) of >0.8 were confirmed for all three immune cell markers.

Nuclear B catenin and membranous E cadherin staining was scored manually by compartment in epithelial cells and weighted histoscores were calculated using the equation below (Kirkegaard et al., 2006): (0×𝑛𝑒𝑔𝑎𝑡𝑖𝑣𝑒 𝑠𝑡𝑎𝑖𝑛𝑖𝑛𝑔 𝑝𝑒𝑟𝑐𝑒𝑛𝑡𝑎𝑔𝑒)+(1 × 𝑤𝑒𝑎𝑘 𝑠𝑡𝑎𝑖𝑛𝑖𝑛𝑔 𝑝𝑒𝑟𝑐𝑒𝑛𝑡𝑎𝑔𝑒) +(2×𝑚𝑜𝑑𝑒𝑟𝑎𝑡𝑒 𝑠𝑡𝑎𝑖𝑛𝑖𝑛𝑔 𝑝𝑒𝑟𝑐𝑒𝑛𝑡𝑎𝑔𝑒) +(3×𝑠𝑡𝑟𝑜𝑛𝑔 𝑠𝑡𝑎𝑖𝑛𝑖𝑛𝑔 𝑝𝑒𝑟𝑐𝑒𝑛𝑡𝑎𝑔𝑒). This produced a range of scores between 0-300, where 0 is no staining and 300 is strong staining. In QuPath tissue classes were annotated (epithelium and lamina propria) using a set brush. To generate a histoscore (H-score), 3 thresholds were manually selected from the DAB channel to represent pixels with weak, moderate, and strong DAB values. Any values below the 1+ threshold will be very weak to negative. Any values between the 1+ and 2+ thresholds will be weak. Any values between the 2+ and 3+ thresholds will be moderate. Any values above the 3+ threshold will be strong. A random trees classifier was built from the cell detection, tissue classification, and selected staining intensities to allow for the generation of a histoscore that represents each core. Manual scoring was performed by a single observer blinded to clinical outcome for 10% of the cores then compared to QuPath generated scores and an intraclass correlation coefficient (ICC) was calculated with ICCs >0.7 for both markers.

Multiplex Immunofluorescence

TMA sections of 2.5-µm thickness were dewaxed and rehydrated by xylene and decreasing concentration of industrial methylated spirit (IMS) by an automated slide stainer (Myreva, Tarragona, Spain). Antigen retrieval was performed by placing the slides in the Antigen Retrieval Buffer (Dewax and HIER buffer, ThermoFisher) and first bringing the buffer to boil at full power for 1 min, kept boiling for 10 min at 20% power, and finally allowed to cool for 15 min. Primary antibody incubation was performed for 30 min at room temperature. Ultravision Quanto Detection System HRP (ThermoFisher) was used for blocking, amplification and secondary antibody incubation. Opal-TSA fluorophores (Akoya Biosciences, Malborough, MA, USA) diluted to 1:400-1:1600 was applied for 10 min. After the final round of antigen retrieval, antibody incubation and Opal-TSA reaction, a final microwaving was performed, followed by application of spectral DAPI and mounting with VectaShield Vibrance (Vector Lab, Newark, CA, USA).

TMA sections of 2.5-µm thickness were dewaxed and antigen-retrieved using Dewax and HIER Buffer-H (epredia) with the PT module (Dako). Primary antibody incubation was performed for 30 minutes at room temperature. The Ultravision Quanto Detection System HRP (epredia) was used for blocking, signal amplification, and secondary antibody incubation. Opal-TSA fluorophores (Akoya Biosciences, Marlborough, MA, USA) were applied at a 1:200 dilution for 10 minutes. Staining was performed using the Link 48 autostainer (DAKO). Following each round of antibody staining, an additional antigen retrieval step was carried out using the PT module. Nuclear counterstaining was performed with Spectral DAPI (1:1000 dilution) for 5 minutes, and slides were mounted using ProLong Diamond Antifade Mountant (ThermoFisher Scientific).

**List of antibodies used in mIF**

| Antibody | Antibody Type | Source (Catalogue number) | Antibody Dilution (ng/ml) | Antigen Retrieval  (pH) | Antibody-Opal Pairing | Staining order |
| --- | --- | --- | --- | --- | --- | --- |
| CD3 | Mouse monoclonal, LN-10 | Leica  (NCL-CD3-565) | 32 | 9 | Opal 570 | 5 |
| CD8 | Mouse monoclonal, C8/144B | Agilent (M7103) | 30 | 8 | Opal 520 | 3 |
| FOXP3 | Mouse monoclonal, 236A/E7 | Abcam (ab20034) | 500 | 9 | Opal 690 | 1 |
| αSMA | Mouse monoclonal, 1A4 | Agilent (M0851) | 11.9 | 9 | Opal 540 | 2 |
| CD68 | Mouse monoclonal, PG-M1 | Agilent (M0876) | 12.5 | 8 | Opal 620 | 4 |
| Pan-cytokeratin | Mouse monoclonal, AE1/AE3 | Thermo (MS-343) | 400 | 6 | Opal 650 | 6 |

Slides were scanned using PhenoImager-HT (Akoya Biosciences), underwent spectral unmixing by InForm using a library of single stained slides. Images were analysed using QuPath (Version 0.5.1).

QuPath analysis for Multiplex Immunofluorescence

Tissues and staining artefacts were visually inspected and excluded from regions of interest (ROI). Tissue segmentation to distinguish the epithelium and the lamina propria was performed by a pixel classifier generated by Random Trees algorithm using Pan-cytokeratin, αSMA and DAPI as detection channels. Object classifiers for CD3^+^, CD8^+^, and FOXP3^+^ cells were generated based on CD3 channel for detection and using intensity threshold. Object classifiers were sequentially applied to detect CD3^+^, CD3^+^CD8^+^, CD3^+^FOXP3^+^ cells. CD68^+^ and αSMA^+^ cells were identified by cell detection using each channel. Cell density (cells/mm^2^) was calculated using number of detections divided by area of ROIs.

Mutational panel:

DNA was prepared by GTRF. FFPE blocks were placed into a microtome and serially sectioned. Two 10-μm curls were taken and stored in a 2 mL DNA/RNA-free tube. Curls were stored in 4℃ until used. Curls were centrifuged briefly before the addition of 300 μL mineral oil and vortexed. They were then heated at 80℃ for 2 min and allowed to cool at RT. A master mix containing 224 μL lysis buffer, 25 μL proteinase K, and 1 μL blue dye (per reaction) was prepared, and 250 μL/sample was added and vortexed. This was followed by centrifugation at 10,000g for 20 sec. The contents were then transferred to Eppendorf tubes and heated at 56℃ for 30 min. This was followed by a 4-hour incubation at 80℃. Then the samples were allowed to cool at RT for 5 min. The aqueous phase of the mixture was then mixed with 10 μL of RNase A, and the samples were incubated for 5 min then centrifuged at full speed for 5 min. The aqueous phase was then promptly transferred to the prepared Maxwell® FFPE Cartridge. The deck tray was set-up with the cartridges for use and the seals removed, and plungers positioned into the cartridge wells. Elution tubes were fitted for each cartridge and 50 μL nuclease-free water was added to each elution tube. The on-screen instructions for the Maxwell® RSC DNA FFPE Kit experiment were followed. After the run was completed, the elution tubes (containing extracted DNA) were retrieved and allowed to come to RT for quantitation, which was performed using a calibrated Qubit™ 4 Fluorochrome. Samples were stored for long-term use at -70℃ or below.

DNA sequencing was performed by the Genomic Innovation Alliance (GIA, UK). The SureSelect XT2 HS2 kit (Part NumbeR: G9983D, Agilent, USA) was used for sample sequencing on the NovaSeq 6000 (Illumina, UK). The SureSelect CancerPlus panel (Design ID: S3225252, Agilent, USA) was used to enrich regions of interest. Resulting library quality and quantity was determined by TapeStation D1000 ScreenTape (Cat. 5067-5582, Agilent, USA). The Next Generation Sequencing analysis pipeline and all steps within it were developed and performed by GIA. In brief, sequencing data were processed, and single nucleotide variation files were generated using the HOLMES pipeline (V1.3., V1.3.1). Bcl2fastq conversion software (V2.19.1.403, V2.20.0.422 on C++; Gerstung et al. 2012, 2014) was used to convert NovaSeq 6000-generated “.cblc” raw data files to FASTQ files and aligned using Burrows-Wheeler Alignment (V0.7.15 as a C program; Li and Durbin 2009). deepSNV/Shearwater (V1.22/V1.1.0, V1.22.0.5/ V1.1.0 as an R package/R wrapper script;Gerstung et al., 2012; Gerstung, Papaemmanuil and Campbell, 2014) was used for calling single nucleotide variants, while Pindel (V0.2.5b8- ww1 as a standalone application; Ye et al. 2009) was used for large 93 insertions/deletions. Once called, variants were annotated using CAVA (V1.2.2.ww1, V1.2.2.ww5 on Python; Münz et al., 2015). Discordant grouping was used to identify structural variation breakpoints using BRASS (V5.3.3-ww10 on C++), and copy-numbers were called using geneCN (V2.1 on Perl and on R). 2.2.7.7 Filtering Strategy The HOLMES pipeline was run in the tumour-only mode on a subset of the INCISE cohort, against a panel of 21 normal tonsil samples. This allowed for germline variants and artefacts to be removed from the INCISE test samples, which generated a purely somatic output

Bulk RNAseq:

Whole human transcriptome bulk RNAseq was carried out on full-section FFPE adenoma samples from the full cohort using by BioClavis Ltd (Biospyder Technologies, Carlsbad, CA, USA) using their proprietary Temp-O-Seq technology.

Unless specified otherwise, all reagents used in Templated Oligo Assay with Sequence Readout (TempO-Seq™) were supplied by BioSpyder (USA). All experiments included human brain reference RNA (BRR) and human universal reference RNA (URR) controls and no sample controls (NSCs) in the same 96-well plate, all run in duplicate. A hybridisation mix containing 2x Annealing Buffer and the detector oligo (DO) pool was prepared as described below. To prevent evaporation during lengthy heated incubations, 6 μL of mineral oil were added to each well of a labelled assay plate. Then 2 μL of Hybridisation Mix was added to each sample well, followed by 2 μL of samples lysed in 2x FFPE Lysis Buffer to the appropriately labelled wells. The plates were sealed, centrifuged at 1000 rpm for 1 min at RT, and placed in a T100 thermal cycler (BioRad). Hybridisation was performed by incubating assay plates at 70℃ for 10 min. The temperature was then reduced to 45℃ by lowering at a rate of 0.5℃/minute, then held at 45℃ for 16 hours. Plates were then cooled to 25℃. To remove unhybridised DOs, a nuclease mix containing 10x Nuclease Buffer, a Nuclease Enzyme, and UltraPure water was prepared, and 24 μL added to each well. The plate was sealed, centrifuged at 1000 rpm for 1 min at RT, and placed in a thermocycler, where it was incubated at 37℃ for 90 minutes. As DOs target adjacent sequences for hybridisation, this allows them to be ligated. A ligation mix consisting of 10x Ligation Buffer, Ligase Enzyme, and UltraPure water was prepared and 24 μL added to each well. The plate was sealed, centrifuged at 1000 rpm for 1 min at RT, and placed in a thermocycler, where it was incubated at 37°C for 1 hour, and then at 80℃ for 15 min. Once ligation was complete, 10μL of ligated product was added to PCR plates containing a proprietary PCR Pre-Mix including indexed primers. The plate was sealed and centrifuged at 1000 rpm for 1 min at RT. The PCR reaction was programmed as follows: 37℃ for 10 min, then 95℃ for 1 min, followed by 35 cycles of 95℃ for 10 sec, then 65℃ for 30 sec, then 68℃ for 30 sec. The optical thermal cycler read the plate after each cycle to measure the amplified product. Finally, the plate was incubated at 68℃ for 2 min and held at 25℃. This was stored at -20℃ or used right away. The amplification was inspected after each run prior to sequencing; a positive amplification curve indicates an amplifiable product that can be sequenced. 20 μL of barcoded PCR products was added to a flip-spin reservoir and purified using the Macherey-Nagel NucleoSpin Gel and PCR Cleanup Kit (Cat. 740609.50, Macherey-Nagel) and quantified by Qubit using the dsDNA kit (Cat. Q32851, ThermoFisher Scientific).

Sequencing was performed on barcoded samples pooled into a single library and run on the Illumina MiniSeq High-Output flowcell. Sequencing reads were demultiplexed by BCL2FASTQ software (Illumina, USA). 2.2.5.10 Analysis of TempO-Seq™ Read Counts Before analysis, sequencing reads in “.FASTQ” format were converted to simple read count tables using BioClavis’ automated pipeline. Sums of read counts from each probe for each sample were averaged, and then normalized by dividing the read count value by the product of the sum of read counts for each probe and the average of probe sums of all samples as shown in the equation below:

𝑅𝑒𝑎𝑑 𝐶𝑜𝑢𝑛𝑡 𝑉𝑎𝑙𝑢𝑒 𝑜𝑓 𝐸𝑎𝑐ℎ 𝑃𝑟𝑜𝑏𝑒 / (𝑆𝑢𝑚 𝑜𝑓 𝑅𝑒𝑎𝑑 𝐶𝑜𝑢𝑛𝑡𝑠 𝑓𝑜𝑟 𝐸𝑎𝑐ℎ 𝑃𝑟𝑜𝑏𝑒 × 𝐴𝑣𝑒𝑟𝑎𝑔𝑒 𝑜𝑓 𝑃𝑟𝑜𝑏𝑒 𝑆𝑢𝑚𝑠 𝑜𝑓 𝐴𝑙𝑙 𝑆𝑎𝑚𝑝𝑙𝑒𝑠)

Quality control and preprocessing was carried out in RStudio (R 4.2.3). Probe variance across all samples was calculated using the *sapply* function. Probes with a variance less than the 25th percentile were removed (n=5640 probes). Samples with a low read count (< 95% CI [<2.28x106 counts]) were identified (*stats* Ver. 4.2.3) and removed (n=110), as well as samples which did not match batch information (n=5). The counts were batch corrected using *ComBat_seq* (sva Ver. 3.46.0). Probe IDs were mapped to gene symbols with duplicated genes collapsed using *MaxMeans* (*WGCNA* Ver. 1.72-1). Normalised counts were generated using quantile normalisation and log2 +1 transformed.

All bulk transcriptomic analysis was performed using R (4.3.3) in RStudio. *DESeq2* (Ver. 1.42.1) was used to perform differential gene expression analysis, and volcano plots were generated using *ggplot2* (Ver. 3.5.0).  *MCPcounter* (v1.2.0) estimated the abundance of tissue infiltrating immune and stromal cell populations using gene expression.

CosMx Experiment:

CosMx™ was performed by Claire Kennedy Dietrich from the Jamieson Spatial Lab

as detailed below. Unless stated otherwise, all reagents are from NanoString.

The FFPE TMA slide was sectioned at 5 μm and placed on a Leica Bond+ slide. The

slide was dried at 37℃ ON followed by baking at 60℃ ON. The slide was then

loaded onto the Lecia Bond automated system for deparaffinisation, followed by

HIER at 100℃ for 15 min in a citrate buffer (pH 6). This was followed by an

incubation in proteinase K (3 μg/mL) for 20 min at 37℃. Slides were then incubated in 0.001% fiducial solution diluted in 2XSSCT for 5 min at RT, and washed sequentially in PBS for 1 min, 10% NBF for 1min, and NBF Stop

buffer for 5 min repeated. The slides were then incubated in NHS-Acetate buffer

for 15 min at RT, followed by two washes in 2XSSC for 5 min. The probe panel

comprising of 6200 genes (including 20 negative control probes) was denatured

at 95 degrees for 2 min before immediately being cooled on ice for at least 1min.

The probes were then mixed with buffer R, RNase inhibitor and DEPC water, and

applied to the slide. Hybridisation was carried out for 17 hours at 37℃ in a

hybridisation oven. The following day the slide was washed twice in a solution of 50% formamide and 50% 4XSSC in a water bath set to 37℃ for 25 min. This was followed by two

washes in 2XSSC for 2 min each, before incubating the slide in DAPI for 15 min at

RT in the dark. The slide was then washed in PBS for 5min before the standard

NanoString antibody panel containing cell membrane marker CD298/B2M,

epithelium marker PanCK, immune marker CD45, and macrophage marker CD68

was prepared with blocking buffer and the slide was incubated for 1 hour at RT

in the dark. The slide was then washed three times in PBS for 5 min each, before

the flow cell was affixed to the slide and 2XSSC was added into the flow cell.

The slide was then processed on the CosMx™ Spatial Molecular Imager (SMI), with

178 fields of view (FOV) selected to ensure the instrument ran within the appropriate time scale. The CosMx™ Instrument then performed 16 imaging cycles, during each cycle 1 out of 4 possible types of reporter fluorophores hybridise to the readout domain of the target probes. After hybridisation, an image was acquired followed by cleavage and removal of the reporter probes. This process was then repeated across all

cycles to capture the complete transcriptomic information. The acquired images

were then decoded into RNA transcripts, which were mapped into a

corresponding segmented cell. The raw files were subsequently exported from the

AtoMx® platform for analysis in R.

CosMx data analysis:

CosMx™ data was quality checked, normalised, underwent dimensionality reduction, integrated, clustered, and analysed by Dr. Assya Legrini. In brief, the dataset was segmented under default settings using AtomMx® and exported as flat text files. The dataset was loaded into R using Seurat (Ver. 5.1.0; Satija et al., 2015; Stuart et al., 2019; Hao et al., 2021, 2024) and cells assigned to the appropriate samples and conditions based on FOV. Morphology-stained images were reviewed on Napari (v.0.4.17) for quality issues including tissue lifting, excessive blurring and cell segmentation. Additionally, cell segmentation was checked post cell type annotation using the package FastReseg (v.1.0.2), only ~2% of cells were flagged, confirming segmentation quality. The final round of QC filtered data based on count (>20), features (>20), size (>300um^2^) and negative proportion (<0.1) metrics. Final metrics were 112 FOVs, 147,449 cells/156,649 original cells and mean count per cell= 263.47.

Log normalization, variable features (selection method = vst, nfeatures = 2000) and scaling of data was all performed using Seurat (v.5.1.0). Batch correction using harmony (v.1.2.4) was run using Patient ID as the grouping variable at theta = 2. Subsequent nearest neighbour graphs (1:30 dimensions), clustering (Louvain method, 0.5 resolution) and UMAP (1:30 dimensions) were run.

mIF clusters were generated from multiplex analysis (above) and incorporated into the Seurat metadata, all downstream analysis was performed using this sub-cohort.

Cell types were identified using semi-supervised methods in InsituType (v.2.0). The Immune-oncology single cell profile was used to identify the major immune/stomal cell types, with 5 unknown clusters generated for additional clusters including epithelial. A flightpath was plotted for all cells, calculating the probability of the cell type being correctly assigned per cluster. All cell types of interest had a confidence of 0.80 and above. The overall macrophage and fibroblast clusters were further subtyped into CD68 +ve and αSMA +ve respectively to mirror the cell types generated in mIF. Epithelial clusters were simply named Epithelial A-E. Annotated cell types were reviewed using the morphology images and location of key cell gene markers. Final cell types were projected on the harmony integrated UMAP. Individual UMAPs were generated for each mIF cluster using the same methods as above. Bar charts showing cell type density percentage per mIF cluster were generated by ggplot2 (v.4.0.0).

Normalization and scale factors was carried out according to the smiDE package (v.0.0.2.04) requirements. Gene overlap metrics was run to ensure cell specific gene comparison, radius was set to 0.05, and filtered for genes with a ratio of 1. A negative binomial regression model was run for mIF cluster per cell types of interest, and a subsequent pairwise comparison was run for differential expression analysis.

Distance metrics were explored between the cell types of interest (CD68 Macrophages, CD4 T cells, CD8 T cells and T regulatory cells) using spatialTIME (v.1.3.4-5). Bivariate Neatest neighbour G(r) using CD68 Macrophages as the anchor cell was performed per patient using 500 permutations, edge correction method set to ‘rs’, and radius = 0-0.5mm at 0.001 increments. Additionally, a mean per cluster was generated for visualisation purposes.
